# Pneumococcal nasopharyngeal carriage in children 2–60 months of age in Novi Sad (Serbia), October 2022–September 2026

**DOI:** 10.64898/2026.11.11.26367886

**Authors:** Vladimir Petrović, Biljana Milosavljević, Milan Djilas, Miloš Marković, Vladimir Vuković, Tatjana Pustahija, Smiljana Rajčević, Snežana Medić, Jelena Djekić Malbaša, Gospava Gvozdenac-Dragičević, Jadranka Lazarević, Milena Zelenović, Renata Bjelić-Novaković, Sanja Trivić, Snežana Kopčanski, Vera Andrić, Mioljub Ristić

## Abstract

**Background:** Streptococcus pneumoniae nasopharyngeal carriage in young children is an important source of transmission and may precede pneumococcal disease. Pneumococcal conjugate vaccines can alter circulating serotypes. Continued surveillance is needed to monitor serotype replacement, vaccine serotype coverage, and antimicrobial resistance.

**Methods:** A prospective surveillance study was conducted in outpatient pediatric practices from October 2022 to September 2026 among children aged 2–60 months in Novi Sad, Serbia. Healthy children attending preventive visits and children with acute respiratory tract infections were enrolled. Nasopharyngeal specimens were collected and cultured for S. pneumoniae. Participant-level analyses used the first eligible specimen per child, while isolate-level analyses included all recovered isolates. Serotyping and antimicrobial susceptibility testing were performed. Theoretical vaccine serotype coverage was estimated for licensed and selected investigational pneumococcal conjugate vaccines, with 23- valent polysaccharide vaccine evaluated separately.

**Results:** Among 3,373 included children, 911 were pneumococcal carriers, giving an overall carriage prevalence of 27.0%. Carriage was higher among children with acute respiratory tract infection than among healthy children and was also associated with daycare attendance, chronic medical conditions, age, season, and surveillance year. In total, 1,312 pneumococcal isolates were recovered, of which 87.9% were typeable. The most frequent serotypes included 11A, 10A, 15A, 15B, 23A, 23B, 19A, and 6B. Theoretical serotype coverage was 8.2% for PCV10, 17.5% for PCV13, 20.0% for PCV15, and 47.3% for PCV20. More than half of typeable isolates were non-PCV20 serotypes. Resistance was most frequent to erythromycin, clindamycin, and tetracycline, while multidrug resistance was observed in 18.1% of isolates. Penicillin G non-susceptibility was uncommon.

**Conclusion:** Pneumococcal carriage in children aged 2–60 months in Novi Sad was associated with age, clinical presentation, season, and surveillance year and involved diverse serotypes. Continued carriage surveillance, linked with invasive-disease and antibiotic-use data, would help determine whether the observed serotype and resistance patterns persist.

## Introduction

Streptococcus pneumoniae is a major bacterial pathogen responsible for respiratory tract infections and invasive pneumococcal disease, including bacteremia, meningitis, and sepsis. Its principal ecological niche is the human nasopharynx, where colonization is frequently asymptomatic. Nasopharyngeal carriage is an essential step in pneumococcal transmission and generally precedes the development of pneumococcal disease (1–6). Carriage is particularly common among young children, who represent an important reservoir for pneumococcal circulation within communities. Reported carriage prevalence varies considerably according to age, geographic setting, social contact patterns, and other epidemiological characteristics (7–11).

Nasopharyngeal carriage surveillance provides valuable information on pneumococcal epidemiology, particularly in settings where laboratory-confirmed invasive pneumococcal disease surveillance is limited. Monitoring carriage prevalence and serotype distribution allows identification of circulating pneumococcal populations, assessment of temporal and seasonal changes, and detection of emerging serotypes and antimicrobial resistance patterns (6,12–14). Although carriage isolates do not necessarily reflect the serotype distribution of invasive disease, they provide important information about the reservoir from which pneumococcal transmission and infection originate (2,5,11).

Pneumococci exhibit substantial capsular diversity, with more than 100 serotypes identified. These serotypes differ in their colonization characteristics, transmission potential, and propensity to cause invasive disease (8,11,14–16).

Pneumococcal populations are also genetically adaptable through natural transformation and acquisition of exogenous genetic material, contributing to the emergence and dissemination of antimicrobial resistance (1,4,17,18).

Consequently, surveillance of circulating serotypes and their antimicrobial susceptibility profiles is important for understanding changes in pneumococcal epidemiology and informing prevention strategies (19–24).

The introduction of pneumococcal conjugate vaccines (PCVs) has substantially reduced vaccine-type carriage and invasive pneumococcal disease. By reducing nasopharyngeal colonization with vaccine-covered serotypes, PCVs also decrease transmission and provide indirect protection to unvaccinated populations (25–36). However, reductions in vaccine-serotype carriage have frequently been accompanied by increases in non-vaccine serotypes, a phenomenon known as serotype replacement (14,27,28,37,38). Recent European carriage studies have identified serotypes such as 11A, 15A, 15B/C, 23A, and 23B among frequently circulating non-PCV13 serotypes (39,40). These changes highlight the importance of continued surveillance and have contributed to the development of higher-valency pneumococcal vaccines intended to broaden serotype coverage (37,41–43).

In Serbia, routine childhood pneumococcal vaccination was introduced into the National Immunization Program on 1 April 2018, initially using PCV10. PCV13 replaced PCV10 in April 2022, with the vaccination schedule changing from 3+1 to 2+1 in January 2023. PCV10 was subsequently reintroduced in January 2024 under the 2+1 schedule (44,45). These program changes occurred during a period in which pneumococcal serotype distribution and antimicrobial resistance patterns could continue to evolve.

Previous pneumococcal carriage data from Serbia were provided by a pilot study conducted in Novi Sad during 2020– 2021 among children aged 24–60 months. That study reported an overall carriage prevalence of 31.7%, with serotypes 15B, 6B, 19F, 11A, 6A, 3, 19A, and 23F among the most frequently identified. Resistance to erythromycin, clindamycin, and tetracycline exceeded 30% (46). These findings provided a local baseline following PCV10 introduction and indicated the need for continued surveillance encompassing a broader pediatric age range and subsequent vaccination-program changes.

The present study was conducted to characterize the epidemiology of nasopharyngeal S. pneumoniae carriage among healthy and sick children aged 2–60 months residing in Novi Sad, Serbia, during four consecutive surveillance years from October 2022 through September 2026. The primary objectives were to estimate carriage prevalence, characterize pneumococcal serotype distribution, identify vaccine-type and epidemiologically relevant non-vaccine-type serotypes, and assess the theoretical proportion of circulating typeable isolates corresponding to serotypes included in licensed and selected investigational higher-valency pneumococcal vaccine formulations. Secondary objectives were to describe seasonal variation in carriage, evaluate temporal changes in serotype distribution, and characterize antimicrobial resistance patterns and their changes over time.

## Materials and methods

### Study Design and Setting

We conducted a prospective, non-interventional epidemiological surveillance study of nasopharyngeal Streptococcus pneumoniae carriage among children aged 2–60 months in Novi Sad, Serbia, from 1 October 2022 to 30 September 2026. Novi Sad is the administrative center of the Autonomous Province of Vojvodina and has approximately 360,925 inhabitants.

The study covered four consecutive surveillance years: Year 1 (October 2022–September 2023), Year 2 (October 2023– September 2024), Year 3 (October 2024–September 2025), and Year 4 (October 2025–September 2026). The surveillance period was completed before database lock and analysis. The final database included all eligible nasopharyngeal specimens collected from 1 October 2022 through 30 September 2026, and no projected or interim data were included.

The study was coordinated by the Faculty of Medicine, University of Novi Sad, in collaboration with the Institute of Public Health of Vojvodina and the Department of Pediatrics of the Primary Health Care Centre Novi Sad. Surveillance encompassed periods during which PCV13 and PCV10 were used in the Serbian National Immunization Program, allowing description of pneumococcal epidemiology during changes in national vaccination policy.

### Study Population and Sampling Strategy

Children aged 2–60 months residing in Novi Sad were recruited consecutively during routine preventive pediatric examinations or consultations for acute respiratory tract infections at participating outpatient pediatric practices.

Seven pediatricians participated in recruitment, data collection, and nasopharyngeal specimen collection. Each provided primary care for approximately 1,500–2,500 children younger than 18 years of age. Training of pediatricians was conducted in September 2022 prior to study initiation.

Eligible children were required to reside in Novi Sad, be aged 2–60 months, attend a participating pediatric practice during surveillance, and have written informed consent provided by a parent or legal guardian. Children who had received systemic antibiotic treatment within seven days before sampling were excluded.

Children were classified into two clinical groups at the time of sampling. Children attending preventive examinations without signs or symptoms of acute infection were defined as healthy. Children presenting with symptoms and clinically diagnosed acute upper or lower respiratory tract infections were defined as sick.

The estimated population of children aged 2–60 months in Novi Sad was approximately 19,000. Based on an expected pneumococcal carriage prevalence of approximately 32% from previous local surveillance (46), the planned sample size was approximately 1,200 nasopharyngeal specimens per surveillance year. This target referred to total specimens, including specimens collected at repeated visits. Participant-level prevalence analyses were restricted to the first eligible specimen from each enrolled child, whereas specimen-level and isolate-level analyses included repeated visits when applicable. Therefore, the numbers of enrolled children, collected specimens, and pneumococcal isolates differ across analyses.

To facilitate evaluation of seasonal variation, approximately 100 specimens were targeted for collection each month, distributed across five age groups (2–11, 12–23, 24–35, 36–47, and 48–60 months), with a target of approximately 20 specimens per age group monthly. Sampling was consecutive within predefined age strata rather than probability-based.

### Analytical Datasets

Two analytical datasets were defined according to the objectives of the study. The participant-level dataset included one observation per child, corresponding to the first eligible nasopharyngeal specimen collected during surveillance. This dataset was used to estimate carriage prevalence and examine associations with demographic characteristics, clinical presentation, vaccination status, season, and surveillance year.

The isolate-level dataset included all recovered S. pneumoniae isolates, including isolates obtained from specimens collected during repeated visits by the same child. It was used to characterize serotype distribution, theoretical serotype coverage, antimicrobial susceptibility, and temporal microbiological changes.

All typeable and non-typeable isolates were included in overall serotype distribution and antimicrobial resistance descriptions. Analyses of theoretical serotype coverage were restricted to typeable isolates. Non-typeable isolates were considered a separate category and were not classified as either vaccine-type or non-vaccine-type.

Because repeated isolates from individual children could contribute correlated observations, isolate-level statistical comparisons and temporal trend tests were considered exploratory. No adjustment for within-child correlation was performed.

### Data Collection

Demographic, clinical, vaccination, and laboratory information were collected using a standardized electronic questionnaire completed by participating pediatricians.

Recorded demographic variables included age, sex, date of sampling, and daycare attendance. Clinical variables included the presence of chronic medical conditions, respiratory symptoms, clinical diagnosis, and pneumococcal vaccination history, including dates of vaccine administration.

Vaccination status was classified as fully vaccinated, partially vaccinated, or unvaccinated according to the information recorded at the first eligible sampling visit for participant-level analyses. Pneumococcal conjugate vaccine vaccination status was determined from documented vaccination records and classified according to the number of PCV doses received before the date of nasopharyngeal sampling. Children were considered fully vaccinated if they had received the age-appropriate complete PCV schedule recommended in the national immunization programme at the time of sampling. Children were considered partially vaccinated if they had received at least one PCV dose but had not completed the age-appropriate recommended schedule by the sampling date. Children were considered unvaccinated if no PCV dose had been documented before sampling. Doses administered on or after the sampling date were not included in the classification. Vaccination histories recorded during subsequent visits were retained in the microbiological database.

Laboratory variables included identification of S. pneumoniae, serotype, and antimicrobial susceptibility results. Data were entered into a dedicated web-based surveillance platform operated by the Institute of Public Health of Vojvodina, which linked epidemiological and laboratory information and incorporated automated quality-control procedures.

Data collection was completed on 30 September 2026 before database lock and analysis. Analyses were conducted in October 2026.

### Nasopharyngeal Specimen Collection and Pneumococcal Identification

Nasopharyngeal specimens were collected according to the recommendations of the World Health Organization Pneumococcal Carriage Working Group (47). Flexible nylon flocked swabs were inserted through the nostril and advanced to the posterior nasopharynx. After collection, specimens were placed in liquid Amies transport medium (Copan, Brescia, Italy) and transported to the Centre for Microbiology of the Institute of Public Health of Vojvodina for processing within six hours.

Specimens were cultured on Columbia agar supplemented with 5% sheep blood and 5 μg/mL gentamicin (ProReady, Kikinda, Serbia) and incubated at 35–37°C in an atmosphere containing 5–10% CO₂. Following 18–24 hours of incubation, presumptive pneumococcal colonies were identified by characteristic morphology, α-hemolysis, optochin susceptibility, and bile solubility testing (47,48). One morphologically typical pneumococcal colony from each positive culture was subcultured and stored in skim milk–tryptone–glucose–glycerol medium (HiMedia, Mumbai, India) at −80°C for subsequent serotype determination.

### Serotype Determination and Molecular Serotyping

Pneumococcal serotyping was performed primarily using the Quellung reaction with type-specific antisera obtained from Statens Serum Institut, Copenhagen, Denmark (49).

Additionally, DNA was extracted from pneumococcal culture isolates according to the Centers for Disease Control and Prevention protocol (50). Species confirmation was performed by amplification of the pneumococcal lytA gene using previously validated primers, with real-time PCR (Eppendorf, Hamburg, Germany) employed when additional confirmation was required (51–53).

Molecular confirmation and additional capsular characterization were performed using sequential multiplex PCR assays based on CDC-recommended protocols and primer schemes (54–56). Serotypes 6A, 6B, and 6C were differentiated using a serotype-specific PCR assay (57). Reference strains ATCC 6305, ATCC 49619, ATCC 6303, and ATCC 700677 were included for quality assurance. PCR products were separated by electrophoresis on 2% agarose gels and visualized using the BioDocAnalyze imaging system.

### Antimicrobial Susceptibility Testing

Antimicrobial susceptibility testing was performed according to EUCAST recommendations using the breakpoint tables applicable at the time of testing, including version 12.0 for 2022 and version 15.0 for 2025 (58,59). Disk diffusion testing was performed using antimicrobial disks (Bio-Rad, Hercules, USA) on Mueller–Hinton agar supplemented with horse blood and β-NAD (Oxoid, Basingstoke, United Kingdom). Oxacillin disks (1 µg) were used to screen for reduced β-lactam susceptibility. Isolates with positive oxacillin screening results underwent penicillin G minimum inhibitory concentration (MIC) testing using gradient diffusion strips (Liofilchem, Roseto degli Abruzzi, Italy) before 2025. From 2025 onward, penicillin G/benzylpenicillin susceptibility among isolates with positive oxacillin screening results was determined using a 1-unit benzylpenicillin disk diffusion test (Bio-Rad, Hercules, USA), according to EUCAST recommendations. Additional ampicillin and ceftriaxone susceptibility testing was performed when indicated according to EUCAST recommendations. Reported antimicrobial susceptibility outcomes included penicillin G non-susceptibility, ampicillin resistance, ceftriaxone resistance, and resistance to erythromycin, clindamycin, trimethoprim–sulfamethoxazole, tetracycline, and levofloxacin. Because susceptibility results were interpreted using the EUCAST breakpoint versions applicable at the time of testing, temporal comparisons were interpreted cautiously, as potential changes in breakpoint criteria across surveillance years could influence resistance classification.

Multidrug resistance (MDR) was defined as resistance to at least one antimicrobial agent in three or more distinct antimicrobial classes. The antimicrobial classes considered were β-lactams, macrolides, lincosamides, tetracyclines, folate pathway inhibitors, and fluoroquinolones. Resistance to multiple agents within the same class was counted only once. Only agent-specific resistance established according to the applicable EUCAST interpretive criteria was considered. Penicillin G, ampicillin, and ceftriaxone were classified within the β-lactam class and were not counted as separate antimicrobial classes.

### Outcome Measures

The primary outcomes were participant-level prevalence of nasopharyngeal S. pneumoniae carriage, pneumococcal serotype distribution, and the proportions of typeable isolates corresponding to serotypes covered by licensed pneumococcal vaccine formulations or classified as non-vaccine types. Theoretical serotype coverage in selected investigational higher-valency pneumococcal conjugate vaccine formulations was also evaluated.

Secondary outcomes were seasonal variation in carriage prevalence, temporal changes in carriage prevalence and serotype distribution, antimicrobial susceptibility patterns, and temporal changes in antimicrobial resistance.

Additional descriptive analyses examined carriage prevalence according to demographic, clinical, and vaccination characteristics and the distribution of vaccine-type isolates according to vaccination status.

### Statistical Analysis

All statistical analyses were conducted using IBM SPSS Statistics version 26 (IBM Corp., Armonk, NY, USA). Statistical tests were two-sided, and p-values below 0.05 were considered statistically significant unless otherwise specified. Continuous variables were assessed for normality using the Shapiro–Wilk test and visual inspection of histograms and quantile–quantile plots. Normally distributed variables were summarized as means and standard deviations, whereas non-normally distributed variables were expressed as medians and interquartile ranges. Categorical variables were reported as counts and percentages.

Comparisons of categorical variables were performed using Pearson’s chi-square test or Fisher’s exact test, as appropriate. Age distributions between healthy and sick children were compared using the Mann–Whitney U test. Variables included in the primary participant-level analyses had complete data, and no missing-data imputation was performed.

Analyses of pneumococcal serotype distribution, theoretical vaccine serotype coverage, and antimicrobial resistance were performed at the isolate level. Repeated isolates from the same child were retained and were not adjusted for within-child correlation; therefore, these analyses were considered exploratory. Non-typeable isolates were included in overall antimicrobial susceptibility analyses but excluded from serotype-specific and vaccine-type analyses. Percentages were calculated using the relevant analysis denominator: typeable isolates for serotype and vaccine-coverage analyses, and isolates with valid susceptibility results for antimicrobial resistance analyses.

Where applicable, temporal trends were assessed using the Cochran–Armitage test for trend, and multiple-comparison adjustments were performed using the Holm method.

### Carriage Prevalence

Nasopharyngeal carriage prevalence was defined as the proportion of children with culture-confirmed S. pneumoniae in their first eligible nasopharyngeal specimen.

Overall prevalence was estimated for the entire study population and separately for healthy and sick children. Additional comparisons were conducted according to age group, sex, daycare attendance, chronic medical conditions, vaccination status, season, and surveillance year.

Ninety-five percent confidence intervals for participant-level prevalence estimates were calculated using the Clopper– Pearson exact binomial method. For graphical presentation of seasonal and monthly prevalence, Wilson score 95% confidence intervals without continuity correction were used.

Age-related and vaccination-status trends were assessed using the Cochran–Armitage test for trend, treating the corresponding categories as ordered.

### Serotype Distribution and Theoretical Serotype Coverage

Overall serotype analyses included typeable and non-typeable isolates, with non-typeable isolates reported separately. Vaccine-type isolates were defined as typeable isolates belonging to serotypes included in the corresponding vaccine formulation. Non-vaccine-type isolates were typeable isolates not included in that formulation.

Theoretical vaccine-serotype coverage was defined as the proportion of typeable carriage isolates whose serotypes are included in each evaluated pneumococcal vaccine formulation. 23-valent polysaccharide vaccine (PPSV23) coverage was evaluated separately and restricted to typeable isolates from children aged 24–60 months. This measure describes serotype match between carriage isolates and vaccine formulations only; it does not estimate vaccine effectiveness or clinical protection.

Theoretical serotype coverage was assessed for licensed PCV10, PCV13, PCV15, PCV20, and PPSV23 formulations, together with selected investigational higher-valency formulations: PCV21/GBP410, PCV21/V116, VAX-24, Pn-MAPS24v, 25vPnC/PF-07872412, IVT PCV-25, Pn-MAPS30plus, and VAX-31. The serotype compositions used to estimate theoretical serotype coverage are provided in Supplementary Table S1 (60–71). Serotype compositions of investigational pneumococcal conjugate vaccine formulations were identified from publicly available sources, including peer-reviewed clinical trial publications, a regulatory product page, a ClinicalTrials.gov record, a manufacturer press release, and an international patent publication. These formulations were used only for theoretical serotype-coverage comparisons and do not imply clinical efficacy.

For licensed PCVs, cumulative and incremental serotype inclusion was evaluated. Proportions were calculated using typeable isolates as the denominator, and 95% confidence intervals were estimated using the Clopper–Pearson exact binomial method.

Overall serotype distributions were compared between isolates recovered from healthy and sick children using Pearson’s chi-square test, with Cramér’s V used to describe association strength. Individual serotypes were compared against all remaining isolates using two-sided Fisher’s exact tests, with unadjusted odds ratios and 95% confidence intervals calculated.

For 48 individual serotype comparisons, including the non-typeable category, p-values were adjusted using the Benjamini–Hochberg procedure to control the false discovery rate.

### Seasonal Variation and Temporal Trends

Seasons were defined as winter (January–March), spring (April–June), summer (July–September), and autumn (October– December).

Seasonal carriage prevalence was calculated using participant-level data for the entire study population and separately for healthy and sick children. Corresponding seasons were pooled across surveillance years to describe recurring seasonal patterns. Differences between seasonal categories were assessed using Pearson’s chi-square test.

Seasonal prevalence was also examined across 16 consecutive seasons. Monthly prevalence was evaluated using two complementary descriptive approaches: pooling corresponding calendar months across the four surveillance years and examining all 48 consecutive surveillance months.

Wilson score 95% confidence intervals without continuity correction were used for the seasonal and monthly graphical analyses. These analyses were descriptive, without fitted linear trend lines or separate inferential tests comparing individual months.

Annual changes in participant-level carriage prevalence were assessed using Pearson’s chi-square test for overall differences and the Cochran–Armitage test for ordered temporal trends. Annual prevalence was examined overall and separately among healthy and sick children.

Temporal changes in serotype distribution, theoretical serotype coverage, and antimicrobial resistance were evaluated annually. Annual proportions were compared using the Cochran–Armitage test for trend where counts were sufficient. Individual-serotype temporal tests were not adjusted for multiple comparisons.

Isolate-level temporal analyses were interpreted as changes in the composition of recovered pneumococcal isolates rather than as independent participant-level changes or causal effects of vaccination-program transitions.

### Antimicrobial Resistance Analysis

Antimicrobial resistance to each agent was expressed as the number and proportion of isolates classified as resistant according to the applicable EUCAST interpretive criteria, with penicillin G non-susceptibility reported separately. Wilson score 95% confidence intervals were calculated for the corresponding proportions.

Resistance patterns were compared according to clinical status, PCV20 vaccine-type classification, and surveillance year using Pearson’s chi-square test or Fisher’s exact test, as appropriate. These analyses were performed at the isolate level, with repeated isolates from some children retained. Consequently, statistical comparisons and temporal patterns were interpreted as exploratory because observations were not necessarily independent.

For the PCV20 vaccine-type versus non-vaccine-type comparison, seven predefined outcomes were evaluated: penicillin G non-susceptibility, resistance to ampicillin, erythromycin, clindamycin, trimethoprim–sulfamethoxazole, and tetracycline, and multidrug resistance (MDR).

P-values were adjusted for these seven comparisons using the Holm–Bonferroni procedure. Both unadjusted and Holm- adjusted p-values were reported, with adjusted p-values below 0.05 considered statistically significant.

Serotype-specific antimicrobial resistance was summarized for the 15 most frequent typeable serotypes, other typeable serotypes, and non-typeable isolates. A descriptive heatmap was used to visualize resistance patterns. No inferential comparisons were performed for the heatmap.

### Vaccination Status Analyses

Participant-level carriage prevalence was compared among fully vaccinated, partially vaccinated, and unvaccinated children using Pearson’s chi-square test, with the Cochran–Armitage test used to assess ordered trends.

The distribution of vaccine-type and non-vaccine-type isolates according to vaccination status was examined descriptively. No inferential comparisons of theoretical serotype coverage across vaccination categories were performed.

Vaccination-status analyses were not interpreted as estimates of vaccine effectiveness.

### Reporting and Ethical Considerations

The study was reported in accordance with the Strengthening the Reporting of Observational Studies in Epidemiology (STROBE) guidelines (72,73).

The study protocol was approved by the Ethics Committee of the Faculty of Medicine, University of Novi Sad (approval number 01-39/1, 24 June 2022). Written informed consent was obtained from the parent or legal guardian of each participating child before enrollment.

All procedures were conducted in accordance with the Declaration of Helsinki and applicable national regulations. Participant confidentiality was maintained, and data were anonymized before analysis.

## Results

### Study Population and Analytical Datasets

During the completed four-year surveillance period, 5,583 children were assessed for eligibility. Of these, 1,857 were excluded because of systemic antibiotic use within seven days before sampling, and 353 otherwise eligible children were not enrolled because written informed consent was not obtained. The participant-level analysis included 3,373 children. In total, 4,945 nasopharyngeal specimens were collected, including 1,572 specimens obtained during repeated visits.

The isolate-level dataset comprised 1,312 isolates recovered from positive cultures, including cultures from repeated visits; 1,153 isolates (87.9%) were typeable and 159 (12.1%) were non-typeable. The participant-level dataset included 911 children with culture-confirmed pneumococcal carriage. The number of isolates exceeded the number of participant-level carriers because isolates from repeated visits were retained in the isolate-level dataset.

The recruitment process, exclusions, and formation of the analytical datasets are illustrated in Figure 1.

**Figure 1.** Study flow diagram

Baseline characteristics of the 3,373 children included in participant-level analyses are shown in Table 1. The median age was 23 months in both healthy and sick children, and the age distribution did not differ significantly between groups (p = 0.323). Sex distribution was also similar, with boys comprising 52.5% of healthy children and 54.0% of sick children (p = 0.382). Daycare attendance was more frequent among sick children than healthy children (59.6% vs. 53.9%; p = 0.001), and chronic medical conditions were also slightly more common among sick children (6.6% vs. 4.9%; p = 0.045). PCV vaccination status differed significantly between groups (p < 0.001): sick children were more often fully vaccinated than healthy children (69.6% vs. 61.7%), whereas partial vaccination and unvaccinated status were more common among healthy children.

**Table 1.** Baseline characteristics of children included in participant-level analyses

| **Characteristic** | **Overall N=3373** | **Healthy n=1730** | **Sick n=1643** | **p-value** |
|---|---|---|---|---|
| Age, months, mean, median (IQR) | 25.79 23 (9-41) | 25.67 23(8-42) | 25.91 23(10-41) | 0.700 |
| Age group, n (%) |  |  |  |  |
| 2–11 months | 987 (29.3) | 521 (30.1) | 466 (28.4) | 0.323 |
| 12–23 months | 737 (21.8) | 367 (21.2) | 370 (22.5) |  |
| 24–35 months | 558 (16.5) | 274 (15.8) | 284 (17.3) |  |
| 36–47 months | 530 (15.7) | 287 (16.6) | 243 (14.8) |  |
| 48–60 months | 561 (16.6) | 281 (16.2) | 280 (17.0) |  |
| Sex, n (%) |  |  |  |  |
| Male | 1796 (53.2) | 908 (52.5) | 888 (54.0) | 0.382 |
| Female | 1577 (46.8) | 822 (47.5) | 755 (46.0) |  |
| Daycare attendance, n (%) |  |  |  |  |
| Yes | 1912 (56.7) | 932 (53.9) | 980 (59.6) | 0.001 |
| No | 1461 (43.3) | 798 (46.1) | 663 (40.4) |  |
| Chronic medical condition, n (%) |  |  |  |  |
| Yes | 193 (5.7) | 85 (4.9) | 108 (6.6) | 0.045 |
| No | 3180 (94.3) | 1645 (95.1) | 1535 (93.4) |  |
| Vaccination status, n (%) |  |  |  |  |
| Fully vaccinated | 2210 (65.5) | 1067 (61.7) | 1143 (69.6) | <0.001 |
| Partially vaccinated | 968 (28.7) | 545 (31.5) | 423 (25.7) |  |
| Unvaccinated | 195 (5.8) | 118 (6.8) | 77 (4.7) |  |

Data are presented as numbers (n) and percentages (%) unless otherwise specified. Age is presented as mean and median (interquartile range [IQR]) in months. Percentages were computed using the total number of children in each analytical group as the denominator: overall population (N = 3,373), healthy children (n = 1,730), and sick children (n = 1,643). Each child was represented once, corresponding to the first eligible sampling visit. The p-values apply only to comparisons between healthy and sick children, not comparisons with the overall population, Categorical variables (age group, sex, daycare attendance, chronic medical conditions and pneumococcal vaccination status) were assessed using Pearson's chi-square test. Age in months was compared using the Mann–Whitney U test. For variables with multiple categories, the p-value represents the overall comparison across all categories rather than a separate test for each category. Statistical significance was defined as p < 0.05; IQR, interquartile range; PCV, pneumococcal conjugate vaccine.

### Nasopharyngeal Carriage Prevalence

Overall, 911 of 3,373 children had culture-confirmed nasopharyngeal carriage of S. pneumoniae, corresponding to a prevalence of 27.0% (95% CI: 25.5–28.5%). Participant-level pneumococcal carriage prevalence by demographic and clinical characteristics is shown in Table 2. Carriage prevalence varied significantly by age group (p < 0.001), increasing from 21.6% among children aged 2–11 months to 32.1% among those aged 24–35 months, before remaining around 29% in older age groups. The trend across age groups was also significant (p for trend < 0.001). Carriage prevalence did not differ significantly by sex, although it was slightly higher among girls than boys (28.0% vs. 26.1%; p = 0.226). Children attending daycare had substantially higher carriage prevalence than those not attending daycare (31.9% vs. 20.7%; p < 0.001). Carriage was also more frequent among children with chronic medical conditions than among those without such conditions (33.7% vs. 26.6%; p = 0.039). In contrast, carriage prevalence did not differ significantly by vaccination status, either when classified as fully, partially, or unvaccinated (p = 0.080; p for trend = 0.734).

**Table 2.**
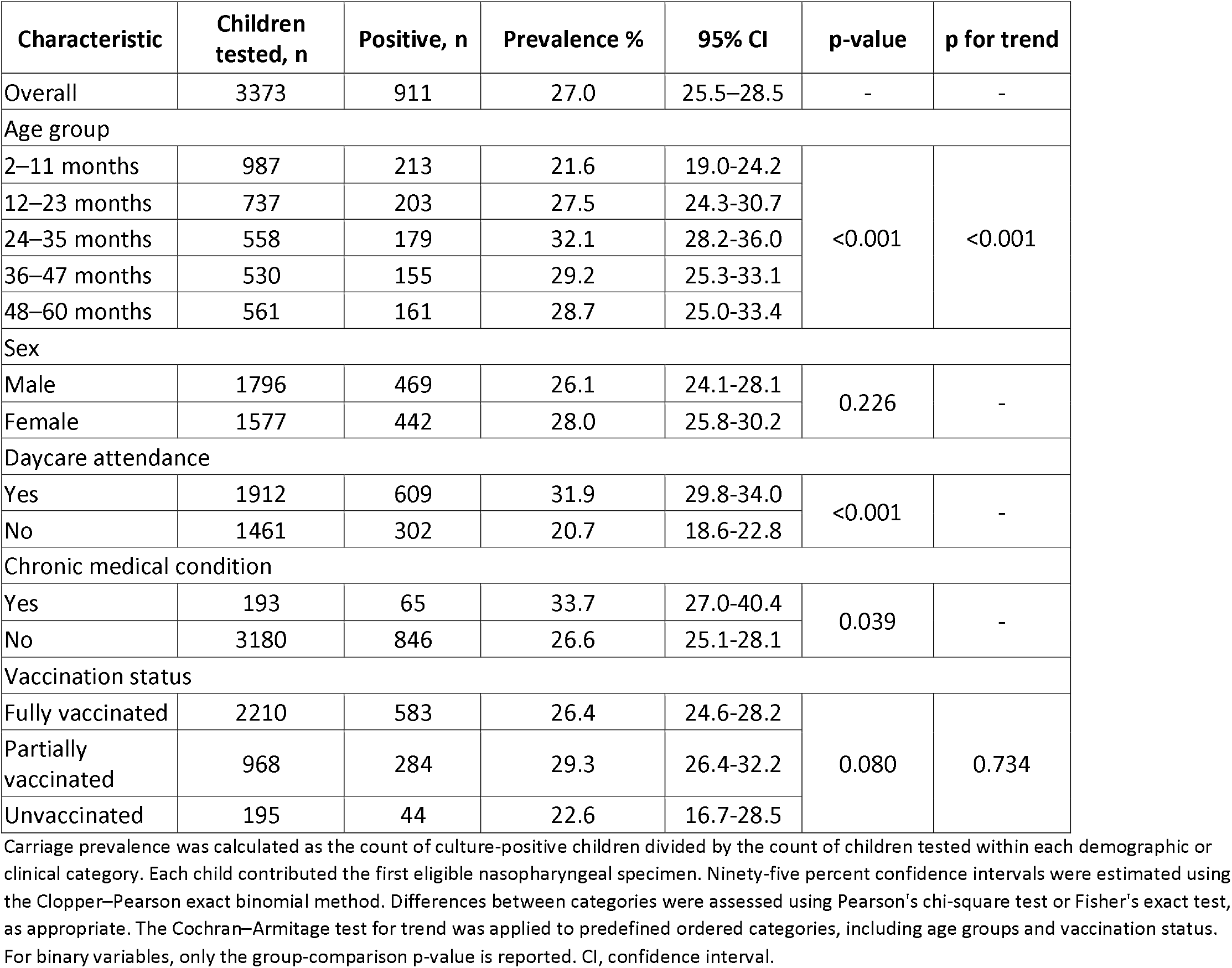
Participant-level pneumococcal carriage prevalence by demographic and clinical characteristics

| Characteristic | Children tested, n | Positive, n | Prevalence % | 95% CI | p-value | p for trend |
|---|---|---|---|---|---|---|
| Overall | 3373 | 911 | 27.0 | 25.5–28.5 | - | - |
| Age group |  |  |  |  |  |  |
| 2–11 months | 987 | 213 | 21.6 | 19.0-24.2 | <0.001 | <0.001 |
| 12–23 months | 737 | 203 | 27.5 | 24.3-30.7 |  |  |
| 24–35 months | 558 | 179 | 32.1 | 28.2-36.0 |  |  |
| 36–47 months | 530 | 155 | 29.2 | 25.3-33.1 |  |  |
| 48–60 months | 561 | 161 | 28.7 | 25.0-33.4 |  |  |
| Sex |  |  |  |  |  |  |
| Male | 1796 | 469 | 26.1 | 24.1-28.1 | 0.226 | - |
| Female | 1577 | 442 | 28.0 | 25.8-30.2 |  |  |
| Daycare attendance |  |  |  |  |  |  |
| Yes | 1912 | 609 | 31.9 | 29.8-34.0 | <0.001 | - |
| No | 1461 | 302 | 20.7 | 18.6-22.8 |  |  |
| Chronic medical condition |  |  |  |  |  |  |
| Yes | 193 | 65 | 33.7 | 27.0-40.4 | 0.039 | - |
| No | 3180 | 846 | 26.6 | 25.1-28.1 |  |  |
| Vaccination status |  |  |  |  |  |  |
| Fully vaccinated | 2210 | 583 | 26.4 | 24.6-28.2 | 0.080 | 0.734 |
| Partially vaccinated | 968 | 284 | 29.3 | 26.4-32.2 |  |  |
| Unvaccinated | 195 | 44 | 22.6 | 16.7-28.5 |  |  |

Carriage prevalence was calculated as the count of culture-positive children divided by the count of children tested within each demographic or clinical category. Each child contributed the first eligible nasopharyngeal specimen. Ninety-five percent confidence intervals were estimated using the Clopper–Pearson exact binomial method. Differences between categories were assessed using Pearson's chi-square test or Fisher's exact test, as appropriate. The Cochran–Armitage test for trend was applied to predefined ordered categories, including age groups and vaccination status. For binary variables, only the group-comparison p-value is reported. CI, confidence interval.

Participant-level pneumococcal carriage prevalence stratified by health status is shown in Table 3. Overall carriage prevalence was higher among sick children than healthy children (32.3% vs. 22.0%). Among healthy children, carriage prevalence differed significantly by age group, increasing from 15.2% in children aged 2–11 months to 28.5% in those aged 24–35 months, and remaining around 26–27% in older age groups (p < 0.001; p for trend < 0.001). In contrast, carriage prevalence among sick children ranged from 28.8% to 35.6% across age groups and did not differ significantly by age group (p = 0.186; p for trend = 0.681). Daycare attendance was associated with higher carriage prevalence in both healthy children (28.5% vs. 14.4%; p < 0.001) and sick children (35.0% vs. 28.2%; p = 0.005). Chronic medical conditions were associated with higher carriage prevalence among sick children (42.6% vs. 31.5%; p = 0.023), but not among healthy children (22.4% vs. 22.0%; p = 1.000). No significant differences in carriage prevalence were observed by sex or vaccination status in either group.

**Table 3.** Participant-level pneumococcal carriage prevalence by demographic and clinical characteristics in Healthy and Sick children

| Characteristic | Healthy |  |  |  |  |  | Sick |  |  |  |  |  |
|---|---|---|---|---|---|---|---|---|---|---|---|---|
|  | Children tested, n | Positive, n | Prevalence % | 95% CI | p-value | p for trend | Children tested, n | Positive, n | Prevalence % | 95% CI | p-value | p for trend |
| Overall | 1730 | 381 | 22.0 | 20.0-24.0 | - | - | 1643 | 530 | 32.3 | 30.0-34.6 | - | - |
| Age group |  |  |  |  |  |  |  |  |  |  |  |  |
| 2–11 months | 521 | 79 | 15.2 | 12.1-18.2 | <0.001 | <0.001 | 466 | 134 | 28.8 | 24.6-32.9 | 0.186 | 0.681 |
| 12–23 months | 367 | 72 | 19.6 | 15.6-23.7 |  |  | 370 | 131 | 35.4 | 30.5-40.3 |  |  |
| 24–35 months | 274 | 78 | 28.5 | 23.1-33.8 |  |  | 284 | 101 | 35.6 | 30.0-41.1 |  |  |
| 36–47 months | 287 | 76 | 26.5 | 21.4-31.6 |  |  | 243 | 79 | 32.5 | 26.6-38.4 |  |  |
| 48–60 months | 281 | 76 | 27.0 | 21.9-32.2 |  |  | 280 | 85 | 30.4 | 25.0-35.7 |  |  |
| Sex |  |  |  |  |  |  |  |  |  |  |  |  |
| Male | 908 | 193 | 21.3 | 18.6-23.9 | 0.452 | - | 888 | 276 | 31.1 | 28.0-34.1 | 0.292 | - |
| Female | 822 | 188 | 22.9 | 20.0-25.7 |  |  | 755 | 254 | 33.6 | 30.3-37.0 |  |  |
| Daycare attendance |  |  |  |  |  |  |  |  |  |  |  |  |
| Yes | 932 | 266 | 28.5 | 25.6-31.4 | <0.001 | - | 980 | 343 | 35.0 | 32.0-38.0 | 0.005 | - |
| No | 798 | 115 | 14.4 | 12.0-16.8 |  |  | 663 | 187 | 28.2 | 24.8-31.6 |  |  |
| Chronic Medical Condition |  |  |  |  |  |  |  |  |  |  |  |  |
| Yes | 85 | 19 | 22.4 | 13.5-31.2 | 1.000 | - | 108 | 46 | 42.6 | 33.3-51.9 | 0.023 | - |
| No | 1645 | 362 | 22.0 | 20.0-24.0 |  |  | 1535 | 484 | 31.5 | 29.2-33.9 |  |  |
| Vaccination status |  |  |  |  |  |  |  |  |  |  |  |  |
| Fully vaccinated | 1067 | 225 | 21.1 | 18.6-23.5 | 0.252 | 0.512 | 1143 | 358 | 31.3 | 28.6-34.0 | 0.164 | 0.523 |
| Partially vaccinated | 545 | 133 | 24.4 | 20.8-28.0 |  |  | 423 | 151 | 35.7 | 31.1-40.3 |  |  |
| Unvaccinated | 118 | 23 | 19.5 | 12.3-26.6 |  |  | 77 | 21 | 27.3 | 17.3-37.2 |  |  |

Carriage prevalence was calculated as the count of culture-positive children divided by the count of children tested within each demographic or clinical category. Each child contributed the first eligible nasopharyngeal specimen. Ninety-five percent confidence intervals were estimated using the Clopper–Pearson exact binomial method. Differences between categories were assessed using Pearson's chi-square test or Fisher's exact test, as appropriate. The Cochran–Armitage test for trend was applied to predefined ordered categories. including age groups and vaccination status. For binary variables. only the group-comparison p-value is reported. Statistical comparisons were performed separately among healthy and sick children. CI – confidence interval.

Supplementary Table S2. shows stratified comparisons of pneumococcal carriage prevalence between healthy and sick children. Overall, carriage prevalence was significantly higher among sick children than healthy children (p < 0.001). This difference was most pronounced in children aged 2–11 months and 12–23 months, where sick children had approximately twice the carriage prevalence observed among healthy children. Significant differences were also observed by sex, daycare attendance, chronic medical condition, fully and partially vaccinated status. In contrast, differences between sick and healthy children were not statistically significant among children aged 24–35, 36–47, or 48–60 months, or among unvaccinated children.

### Pneumococcal Serotype Distribution

Among 1,312 pneumococcal isolates, 1,153 (87.9%) were typeable and 159 (12.1%) were non-typeable. The most frequently identified serotypes were 11A (8.5%), 10A (7.2%), 15A (7.1%), 15B (6.3%), 23A (6.3%), 23B (6.2%), 19A (4.9%), and 6B (4.8%). The distribution of the 15 most frequent serotypes (85.7% of all typeable isolates and 73.6% of all isolates), other typeable serotypes, and non-typeable isolates is presented in Table 4.

**Table 4.**
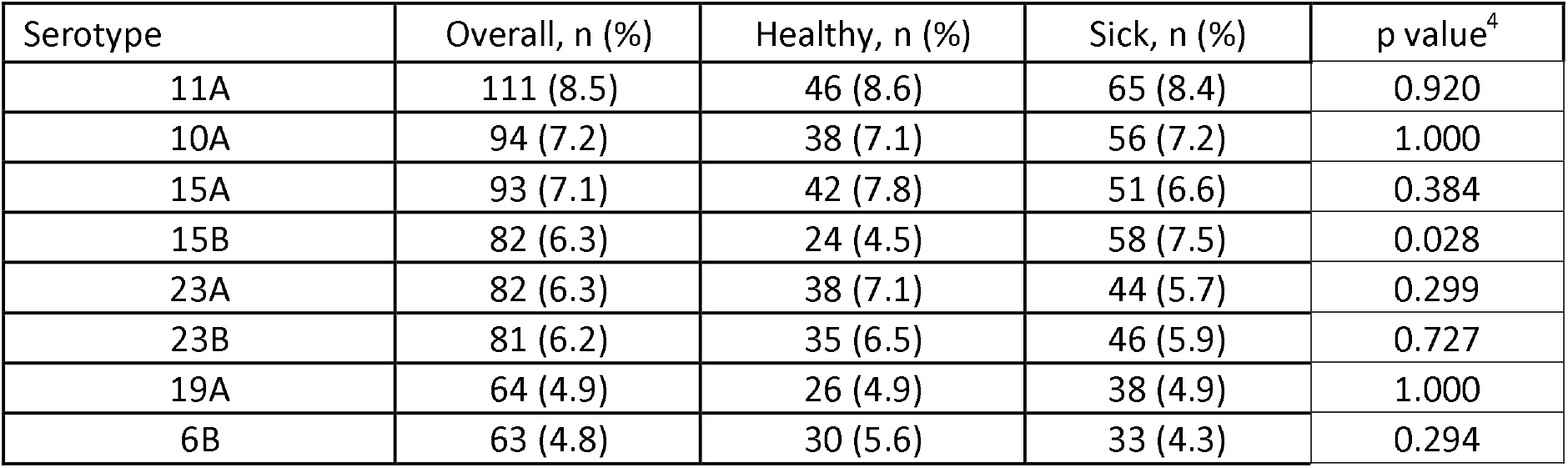
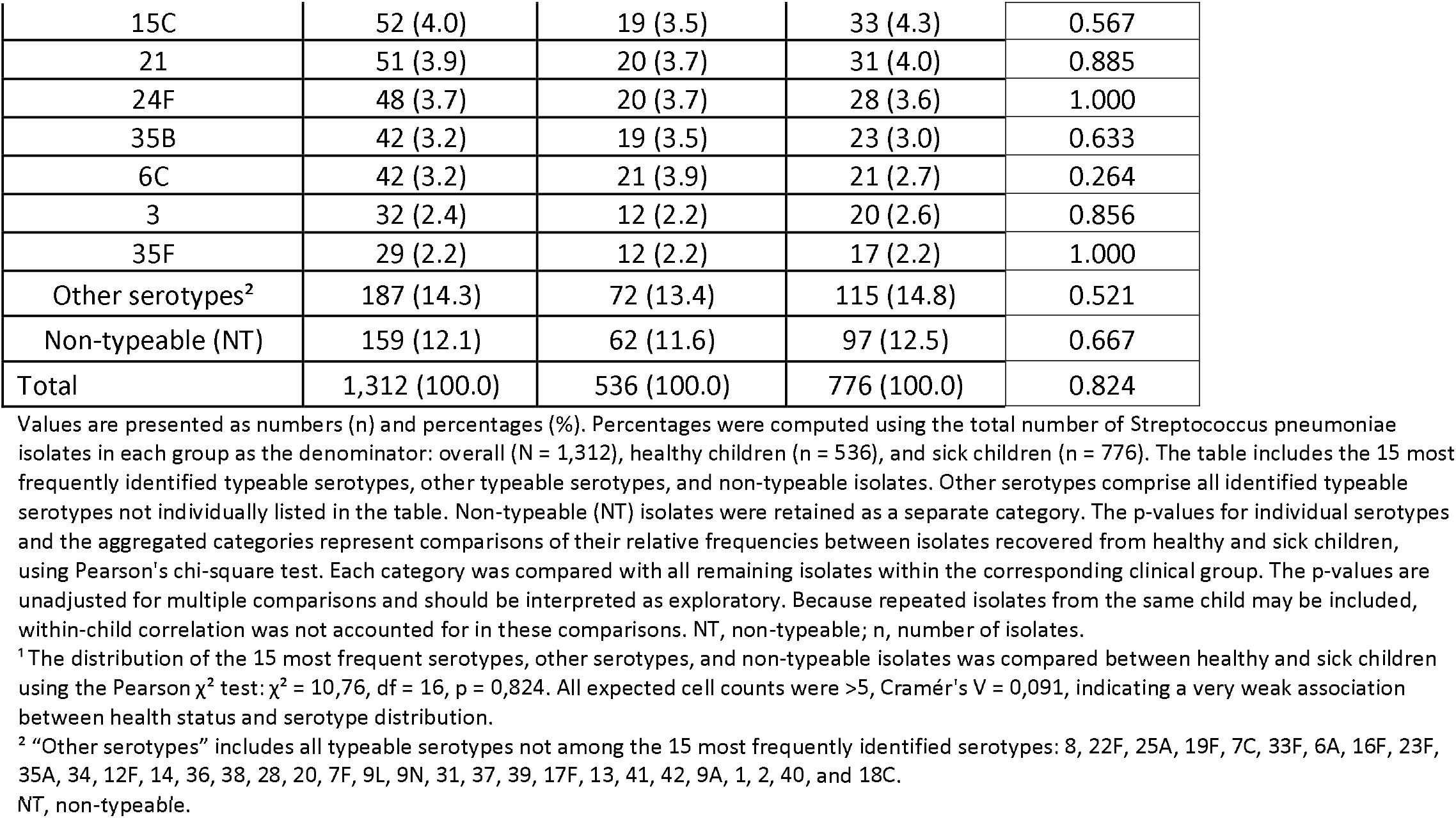
Distribution of the 15 most frequent Streptococcus pneumoniae serotypes among healthy and sick children

The overall distribution of these serotype categories did not differ significantly between isolates recovered from healthy and sick children (p = 0.824). Serotype 15B was more frequently identified among isolates from sick than healthy children (7.5% vs. 4.5%; unadjusted p = 0.028). However, this difference did not remain statistically significant after adjustment for multiple comparisons. The complete distribution of identified serotypes, including less frequent serotypes, and the results of individual serotype comparisons with multiple-testing adjustment are provided in Supplementary Table S3.

### Theoretical Serotype Coverage in Licensed Pneumococcal Vaccines

Among 1,153 typeable isolates, 94 (8.2%) belonged to serotypes included in PCV10, 202 (17.5%) in PCV13, 231 (20.0%) in PCV15, and 545 (47.3%) in PCV20 (Table 5). Consequently, 608 isolates (52.7%) belonged to serotypes not included in PCV20.

**Table 5.** Serotype inclusion of pneumococcal isolates in licensed pneumococcal conjugate vaccine formulations

| Vaccine formulation | Included serotype isolates. n (%) | 95% CI | NVT isolates n (%) |
|---|---|---|---|
| PCV10 | 94 (8.2) | 6.6-9.8 | 1059 (91.8) |
| PCV13 | 202 (17.5) | 15.3-19.7 | 951 (82.5) |
| PCV15 | 231 (20.0) | 17.7-22.3 | 922 (80.0) |
| PCV20 | 545 (47.3) | 44.8-50.6 | 608 (52.7) |

Values are presented as numbers (n) and percentages (%). Theoretical vaccine-serotype inclusion was calculated among 1,153 typeable Streptococcus pneumoniae isolates recovered during the surveillance period. Included serotype isolates represent isolates belonging to serotypes contained in the corresponding pneumococcal conjugate vaccine (PCV) formulation. Non-vaccine-type (NVT) isolates represent typeable isolates with serotypes not included in the corresponding vaccine formulation. Percentages were computed using the total number of typeable isolates (N = 1,153) as the denominator for each vaccine formulation. Ninety-five percent confidence intervals (95% CI) for vaccine-serotype inclusion were estimated using the Clopper–Pearson exact binomial method. Non-typeable isolates (n = 159) were excluded from both vaccine-type and NVT categories. Vaccine-serotype inclusion was assessed cumulatively for each formulation and represents theoretical serotype coverage rather than vaccine effectiveness or protection against pneumococcal disease. PCV10, 10-valent pneumococcal conjugate vaccine; PCV13, 13-valent pneumococcal conjugate vaccine; PCV15, 15-valent pneumococcal conjugate vaccine; PCV20, 20-valent pneumococcal conjugate vaccine; NVT, non-vaccine type; CI, confidence interval.

Separately, among children aged 24–60 months, PPSV23 theoretical serotype coverage was 47.1%: 319 of 678 typeable isolates in this age group belonged to serotypes included in PPSV23. Six serotypes included in PPSV23 but not in the licensed PCV10/13/15/20 formulations were classified as non-vaccine types for PCV-specific calculations. Because PPSV23 is not routinely used in younger children, this analysis was restricted to children aged 24–60 months.

The greatest incremental increase in theoretical serotype coverage occurred between PCV15 and PCV20 and the smallest occurred between PCV13 and PCV15. The corresponding cumulative and incremental serotype categories are presented in Table 6.

**Table 6.** Incremental serotype inclusion across licensed PCV formulations

| Category | Serotypes included | Isolates n (%) |
|---|---|---|
| PCV10 serotypes | 1, 4, 5, 9V, 14, 18C, 19F, 23F, 6B, 7F | 94 (8.2) |
| Additional PCV13-non-PCV10 serotypes (PCV13-PCV10) | 3, 6A, 19A | 108 (9.3) |
| Additional PCV15-non-PCV13 serotypes (PCV15-PCV13) | 22F, 33F | 29 (2.5) |
| Additional PCV20-non-PCV15 serotypes (PCV20-PCV15) | 8, 10A, 11A, 12F, 15B | 314 (27.3) |
| Non-PCV20 serotypes | all others | 608 (52.7) |

Values are presented as numbers (n) and percentages (%). Percentages were computed using all 1,153 typeable Streptococcus pneumoniae isolates as the denominator. Serotypes were classified into mutually exclusive categories according to their inclusion in licensed pneumococcal conjugate vaccine (PCV) formulations. The PCV10 category includes serotypes represented in PCV10; the PCV13–PCV10, PCV15–PCV13, and PCV20–PCV15 categories represent additional serotypes included in each successive higher-valency formulation but absent from the preceding formulation. Non-PCV20 serotypes comprise all remaining typeable serotypes not included in PCV20. Each isolate was assigned to one category only; therefore, the categories are mutually exclusive and their percentages sum to 100%. Non-typeable isolates (n = 159) were excluded from the analysis. The proportions represent the distribution of recovered pneumococcal isolates according to theoretical vaccine-serotype inclusion and should not be taken as vaccine effectiveness or protection against pneumococcal disease. PCV10, 10-valent pneumococcal conjugate vaccine; PCV13, 13-valent pneumococcal conjugate vaccine; PCV15, 15-valent pneumococcal conjugate vaccine; PCV20, 20-valent pneumococcal conjugate vaccine.

Theoretical serotype coverage was also examined according to clinical status (Table 7). Theoretical serotype coverage was similar between healthy and sick children for PCV10, PCV13, and PCV15, while PCV20 coverage was numerically higher among isolates from sick children than healthy children, although this difference was not statistically significant.

**Table 7.** Theoretical serotype coverage of pneumococcal isolates in licensed pneumococcal conjugate vaccine formulations by clinical status of children

| Vaccine formulation | Healthy children n=474 |  |  | Sick children n=679 |  |  | P value |
|---|---|---|---|---|---|---|---|
|  | Covered serotype isolates, n (%) | 95% CI | NVT isolates n (%) | Covered serotype isolates, n (%) | 95% CI | NVT isolates n (%) |  |
| PCV10 | 42 (8.9) | 6.3-11.4 | 432 (91.1) | 52 (7.7) | 5.7-9.7 | 627 (92.3) | 0.463 |
| PCV13 | 84 (17.7) | 14.3-21.2 | 390 (82.3) | 118 (17.4) | 14.5-20.2 | 561 (82.6) | 0.880 |
| PCV15 | 91 (19.2) | 15.7-22.7 | 383 (80.8) | 140 (20.6) | 17.6-23.7 | 539 (79.4) | 0.553 |
| PCV20 | 211 (44.5) | 40.0-49.0 | 263 (55.5) | 334 (49.2) | 45.4-53.0 | 345 (50.8) | 0.118 |

Values are presented as numbers (n) and percentages (%). The analysis included 1,153 typeable Streptococcus pneumoniae isolates recovered from healthy children (n = 474) and sick children (n = 679). Percentages were calculated separately within each clinical group, using the corresponding number of typeable isolates as the denominator. Included serotype isolates represent isolates belonging to serotypes contained in each pneumococcal conjugate vaccine (PCV) formulation. Non-vaccine-type (NVT) isolates represent typeable isolates with serotypes not included in the corresponding formulation. Ninety-five percent confidence intervals (95% CI) for vaccine-serotype inclusion were estimated using the Clopper–Pearson exact binomial method. The p-values indicate comparisons of vaccine-serotype inclusion between isolates recovered from healthy and sick children for each vaccine formulation, using Pearson's chi-square test or Fisher's exact test, as appropriate. Non-typeable isolates (n = 159; healthy, n = 62; sick, n = 97) were excluded. Vaccine-serotype inclusion was assessed cumulatively for each formulation. Because repeated isolates from the same child may be included, the comparisons are exploratory and do not account for within-child correlation. Theoretical vaccine-serotype inclusion should not be taken as vaccine effectiveness or protection against pneumococcal disease. PCV, pneumococcal conjugate vaccine; NVT, non-vaccine type; CI, confidence interval.

These findings describe the correspondence between circulating pneumococcal serotypes and vaccine formulations rather than observed vaccine effectiveness.

### Theoretical Serotype Coverage in Investigational Higher-Valency Pneumococcal Vaccines

Full results for all evaluated investigational formulations are shown in Table 8. Among 1,153 typeable isolates, the highest overall theoretical coverage was observed for Pn-MAPS30plus (89.0%), followed by 25vPnC/PF-07872412 (77.4%) and VAX-31 (73.4%); corresponding coverage was 68.8% for PCV21/V116 and 67.9% for IVT PCV-25. Pn- MAPS30plus also covered the largest proportion of non-PCV20 serotypes and left the fewest typeable isolates uncovered. These estimates are descriptive theoretical serotype-inclusion measures and do not imply comparative vaccine effectiveness or protection against pneumococcal disease.

**Table 8.** Theoretical serotype coverage of investigational higher-valency pneumococcal conjugate vaccine formulations among Streptococcus pneumoniae isolates, including additional

| Investigational formulation | All isolates covered, n (%) | non PCV20 vaccine type isolates covered, n (%) | Remaining non-covered isolates, n (%) |
|---|---|---|---|
| PCV21/GBP410 | 548 (47.5) | 3 (0.3) | 605 (52.5) |
| PCV21/V116 | 793 (68.8) | 421 (36.5) | 360 (31.2) |
| VAX-24 | 553 (48.0) | 8 (0.7) | 600 (52.0) |
| Pn-MAPS24v | 553 (48.0) | 8 (0.7) | 600 (52.0) |
| 25vPnC/PF-07872412 | 892 (77.4) | 347 (30.1) | 261 (22.6) |
| IVT PCV-25 | 783 (67.9) | 238 (20.6) | 370 (32.1) |
| Pn-MAPS30plus | 1,026 (89.0) | 481 (41.7) | 127 (11.0) |
| VAX-31 | 846 (73.4) | 301 (26.1) | 307 (26.6) |

Values are presented as n (%) of the 1,153 typeable pneumococcal isolates. Non-typeable isolates (n = 159) were excluded. “All isolates included” represents the number and proportion of typeable isolates with serotypes included in each investigational vaccine formulation. “PCV20-NVT isolates included” represents isolates belonging to serotypes absent from PCV20 but included in the investigational formulation. This measure should not be taken as net incremental coverage over PCV20 because investigational formulations may omit some PCV20 serotypes. “Remaining non-included isolates” represents typeable isolates with serotypes absent from the corresponding investigational formulation. Percentages are descriptive theoretical serotype-inclusion estimates and do not imply vaccine effectiveness.

### Seasonal Variation in Nasopharyngeal Carriage

Pneumococcal carriage prevalence showed seasonal variation (Table 9). Overall carriage prevalence was highest in autumn and winter, at 31.0% and 30.9%, respectively, followed by spring at 29.0%, and was lowest in summer at 16.5%. Seasonal variation was statistically significant in the overall population (p < 0.001), as well as among healthy children and sick children separately (both p < 0.001). Within each season, carriage prevalence was numerically higher among sick children than among healthy children: 35.6% versus 26.1% in autumn, 37.5% versus 24.8% in winter, 34.8% versus 23.4% in spring, and 19.7% versus 13.7% in summer. These within-season differences were not formally tested.

**Table 9.** Seasonal variation in participant-level pneumococcal carriage prevalence, overall and by clinical presentation

| **Season** | **Overall** |  |  |  |  | **Healthy** |  |  |  |  | **Sick** |  |  |  |  |
|---|---|---|---|---|---|---|---|---|---|---|---|---|---|---|---|
|  | **Children tested, n** | **Positive, n** | **Prevalence %** | **95% CI** | **p-value** | **Children tested, n** | **Positive, n** | **Prevalence %** | **95% CI** | **p-value** | **Children tested, n** | **Positive, n** | **Prevalence %** | **95% CI** | **p-value** |
| Autumn | 953 | 295 | 31.0 | 28.0-34.0 | <0.001 | 464 | 121 | 26.1 | 22.1-30.3 | <0.001 | 489 | 174 | 35.6 | 31.3-40.0 | <0.001 |
| Winter | 777 | 240 | 30.9 | 27.7-34.3 |  | 404 | 100 | 24.8 | 20.6-29.3 |  | 373 | 140 | 37.5 | 32.6-42.7 |  |
| Spring | 842 | 244 | 29.0 | 25.9-32.2 |  | 431 | 101 | 23.4 | 19.5-27.7 |  | 411 | 143 | 34.8 | 30.2-39.6 |  |
| Summer | 801 | 132 | 16.5 | 14.0-19.2 |  | 431 | 59 | 13.7 | 10.6-17.3 |  | 370 | 73 | 19.7 | 15.8-24.2 |  |

Seasonal carriage prevalence was calculated using the first eligible nasopharyngeal specimen per child. Seasonal denominators were 953, 777, 842, and 801 children for autumn, winter, spring, and summer, respectively. Within each season, the denominators for healthy and sick children are provided separately. Ninety-five percent confidence intervals were estimated using the Clopper–Pearson exact binomial method. P-values represent Pearson's chi-square comparisons across the four seasons within the overall, healthy, and sick populations. Seasons were treated as nominal categories; no linear trend test was performed.

Monthly aggregation showed broadly similar seasonal fluctuation, with elevated prevalence during the colder months and early spring and lower prevalence during summer (Figure 2). Similar patterns were observed among healthy and sick children, with carriage prevalence generally higher among sick children across most months.

**Figure 2.** Pneumococcal carriage prevalence by months and clinical status Monthly prevalence of nasopharyngeal Streptococcus pneumoniae carriage among overall, healthy, and sick children in Novi Sad, Serbia, during 2022–2026. Bars represent carriage prevalence (%), and error bars indicate 95% confidence intervals calculated using the Wilson score method without continuity correction. Monthly data were aggregated across the four surveillance years.

Analysis of 16 consecutive seasons demonstrated temporal variability in pneumococcal carriage prevalence across surveillance years (Supplementary Figure S1). Seasonal prevalence fluctuated within and between years, without a consistent monotonic increase or decrease. Carriage prevalence was generally higher among sick children than among healthy children, although some seasonal estimates had wide and overlapping 95% confidence intervals.

Analysis of 48 consecutive surveillance months demonstrated marked temporal variability in pneumococcal carriage prevalence across the study period (Supplementary Figure S2). Monthly prevalence declined during 2024 and increased again during the 2025/2026 respiratory season. This pattern was observed in both healthy and sick children and was generally more evident among sick children, although several monthly estimates had wide 95% confidence intervals.

### Temporal Changes in Carriage Prevalence

Overall pneumococcal carriage prevalence differed significantly across surveillance years, decreasing from 32.1% in Year 1 to 29.1% in Year 2 and 16.3% in Year 3, before increasing to 24.9% in Year 4 (Supplementary Table S4). Although the Cochran–Armitage test indicated a significant ordered trend, the annual pattern was not monotonic; therefore, this result should be interpreted as evidence of overall temporal variation rather than a sustained year-on-year decline.

This temporal pattern was also evident when stratified by clinical status (Supplementary Table S5). Among healthy children, prevalence was 25.9%, 25.2%, 13.1%, and 19.0% in Years 1–4, respectively, while among sick children it was 38.7%, 33.3%, 19.6%, and 31.4%. Prevalence was significantly higher among sick than healthy children in each surveillance year. Differences across surveillance years and tests for trend were statistically significant in both groups.

However, the increase in Year 4 indicates that the reduction in carriage prevalence was not sustained uniformly throughout surveillance, consistent with the seasonal and monthly fluctuations observed during the study period.

### Temporal Changes in Pneumococcal Serotype Distribution

Pneumococcal serotype distribution varied across the four surveillance years (Table 10, Figure 3). Descriptively, Figure 3 shows shifts in the relative contribution of both common and infrequent typeable serotypes by surveillance year. In Table 10, unadjusted trend analyses among the 15 most frequently identified serotypes showed decreasing trends for serotypes 23A and 6B and increasing trends for serotypes 23B, 21, and 35F. The clearest opposing patterns were observed for serotypes 23A, which declined from 9.1% in Year 1 to 3.3% in Year 4, and 23B, which increased from 4.4% to 12.4% over the same period; both had unadjusted p for trend < 0.001. Other serotypes exhibited variable annual distributions without consistent temporal trends.

**Figure 3.** Temporal change in all isolated pneumococcal serotypes Grouped bars represent the percentage of each pneumococcal serotype in typeable isolates recovered during four surveillance years: Year 1 (October 2022–September 2023; n = 363), Year 2 (October 2023–September 2024; n = 329), Year 3 (October 2024–September 2025; n = 187), and Year 4 (October 2025–September 2026; n = 274). Percentages were calculated separately for each surveillance year using the corresponding number of typeable isolates as the denominator. The figure includes both frequently and infrequently identified serotypes. Non-typeable isolates (n = 159) were excluded. Repeated isolates from individual children may contribute to the annual distributions. The figure is descriptive, and no statistical comparisons are displayed. Temporal trends for the 15 most frequently identified serotypes are reported separately in the corresponding table.

**Table 10.** Annual distribution of major pneumococcal serotypes

| Serotype | Year 1 |  | Year 2 |  | Year 3 |  | Year 4 |  | P trend |
|---|---|---|---|---|---|---|---|---|---|
|  | Serotype isolates, n | % | Serotype isolates, n | % | Serotype isolates, n | % | Serotype isolates, n | % |  |
| 11A | 36 | 9.9 | 34 | 10.3 | 18 | 9.6 | 23 | 8.4 | 0.498 |
| 10A | 29 | 8.0 | 34 | 10.3 | 16 | 8.6 | 15 | 5.5 | 0.212 |
| 15A | 29 | 8.0 | 33 | 10.0 | 10 | 5.3 | 21 | 7.7 | 0.510 |
| 15B | 31 | 8.5 | 23 | 7.0 | 16 | 8.6 | 12 | 4.4 | 0.082 |
| 23A | 33 | 9.1 | 31 | 9.4 | 9 | 4.8 | 9 | 3.3 | <0.001 |
| 23B | 16 | 4.4 | 16 | 4.9 | 15 | 8.0 | 34 | 12.4 | <0.001 |
| 19A | 23 | 6.3 | 17 | 5.2 | 10 | 5.3 | 14 | 5.1 | 0.528 |
| 6B | 38 | 10.5 | 19 | 5.8 | 2 | 1.1 | 4 | 1.5 | <0.001 |
| 15C | 12 | 3.3 | 18 | 5.5 | 13 | 7.0 | 9 | 3.3 | 0.784 |
| 21 | 6 | 1.7 | 12 | 3.6 | 16 | 8.6 | 17 | 6.2 | <0.001 |
| 24F | 4 | 1.1 | 23 | 7.0 | 10 | 5.3 | 11 | 4.0 | 0.109 |
| 35B | 7 | 1.9 | 16 | 4.9 | 4 | 2.1 | 15 | 5.5 | 0.066 |
| 6C | 24 | 6.6 | 3 | 0.9 | 3 | 1.6 | 12 | 4.4 | 0.149 |
| 3 | 13 | 3.6 | 5 | 1.5 | 6 | 3.2 | 8 | 2.9 | 0.836 |
| 35F | 4 | 1.1 | 9 | 2.7 | 6 | 3.2 | 10 | 3.6 | 0.039 |

Values are presented as numbers (n) and percentages (%). Percentages were computed using the total number of typeable Streptococcus pneumoniae isolates recovered during each surveillance year as the denominator: Year 1 (October 2022–September 2023; n = 363), Year 2 (October 2023–September 2024; n = 329), Year 3 (October 2024–September 2025; n = 187), and Year 4 (October 2025–September 2026; n = 274). The table presents the 15 most frequently identified typeable serotypes across the four-year surveillance period. P for trend values were computed using the Cochran–Armitage test for trend to assess changes in the percentage of each serotype in typeable isolates across the four ordered surveillance years. Each serotype was compared with all other typeable serotypes combined. Because repeated isolates from the same child may be included, the trend analyses are exploratory and do not account for within-child correlation. P-values were not adjusted for multiple comparisons and should be interpreted cautiously. Statistical significance was defined as p < 0.05.

The theoretical serotype coverage of circulating typeable pneumococcal isolates by licensed pneumococcal conjugate vaccine formulations varied across surveillance years (Supplementary Table S6). Coverage declined over time for all formulations: PCV10 from 17.1% in Year 1 to 2.6% in Year 4, PCV13 from 29.5% to 10.6%, PCV15 from 32.0% to 13.9%, and PCV20 from 59.0% to 37.2%. PCV20 provided the highest theoretical coverage in each surveillance year, but its coverage also decreased over the study period. Differences across years and unadjusted tests for linear trend were statistically significant for all formulations. Vaccine-type categories were cumulative and not mutually exclusive.

### Antimicrobial Resistance Patterns

Antimicrobial resistance patterns among 1,312 pneumococcal isolates are shown in Table 11. Agent-specific β-lactam non-susceptibility or resistance was uncommon: penicillin G non-susceptibility was detected in 17 isolates (1.3%; 95% CI: 0.8–2.1), ampicillin resistance in 73 isolates (5.6%; 95% CI: 4.5–6.9), and ceftriaxone resistance in only one isolate (0.1%; 95% CI: 0.01–0.4). Among non-β-lactam agents, resistance was most common to erythromycin (27.1%; 95% CI: 24.8–29.6), followed by clindamycin (19.9%; 95% CI: 17.8–22.1), tetracycline (18.1%; 95% CI: 16.2–20.3), and trimethoprim– sulfamethoxazole (10.4%; 95% CI: 8.8–12.1). No levofloxacin resistance was detected. Multidrug resistance was identified in 18.1% of isolates.

**Table 11.** Antimicrobial non-susceptibility and resistance among pneumococcal isolates

| **Antimicrobial agent** | **Isolates tested, n** | **Non-susceptible/resistant, n** | **Non-susceptible/resistant, %** | **95% CI** |
|---|---|---|---|---|
| Penicillin G | 1312 | 17 | 1.3 | 0.8-2.1 |
| Ampicillin | 1312 | 73 | 5.6 | 4.5-6.9 |
| Ceftriaxone | 1312 | 1 | 0.1 | 0.01-0.4 |
| Erythromycin | 1312 | 356 | 27.1 | 24.8-29.6 |
| Clindamycin | 1312 | 261 | 19.9 | 17.8-22.1 |
| Trimethoprim-sulfamethoxazole | 1312 | 136 | 10.4 | 8.8-12.1 |
| Tetracycline | 1312 | 238 | 18.1 | 16.2-20.3 |
| Levofloxacin | 1312 | 0 | 0.0 | 0.0-0.3 |
| MDR | 1312 | 238 | 18.1 | 16.2-20.3 |

Values are counts and percentages of isolates with antimicrobial non-susceptibility or resistance, with 95% confidence intervals calculated using the Wilson score method. Both typeable and non-typeable isolates were included. Penicillin G results refer to non-susceptibility rather than confirmed resistance. Confirmed antimicrobial resistance was defined using applicable EUCAST clinical breakpoints. MDR was defined as resistance to at least one antimicrobial agent in three or more distinct antimicrobial classes. CI, confidence interval; MDR, multidrug resistance.

Resistance patterns were also examined according to clinical status (Supplementary Table S7). Resistance proportions were similar among isolates from healthy and sick children, with no statistically significant differences observed for penicillin G, ampicillin, erythromycin, clindamycin, trimethoprim–sulfamethoxazole, tetracycline, or multidrug resistance. Erythromycin resistance was frequent in both groups (27.4% among isolates from healthy children and 26.9% among isolates from sick children), and multidrug resistance was observed in 18.3% and 18.0% of isolates, respectively. P-values were unadjusted.

Resistance patterns were compared between PCV20 vaccine-type (VT) and non-vaccine-type (NVT) isolates (Supplementary Table S8). Ampicillin resistance was significantly higher among VT than NVT isolates after Holm adjustment (6.1% vs 2.1%; unadjusted p < 0.001; Holm-adjusted p = 0.005). No other resistance outcome, including penicillin G non-susceptibility, erythromycin, clindamycin, trimethoprim–sulfamethoxazole, tetracycline, or multidrug resistance, differed significantly after adjustment. Complete unadjusted and Holm-adjusted comparisons are presented in Supplementary Table S8. Comparisons were restricted to typeable isolates.

### Temporal Changes in Antimicrobial Resistance

Temporal trends in antimicrobial resistance are shown in Table 12 and Figure 4. Overall, resistance patterns varied significantly across study years. Declining trends were observed for ampicillin resistance, erythromycin resistance, clindamycin resistance, tetracycline resistance, and multidrug resistance. Clindamycin resistance decreased from 24.2% in Year 1 to 13.9% in Year 4, tetracycline resistance from 22.1% to 11.3%, and multidrug resistance from 22.6% to 11.6%. Erythromycin resistance remained among the most frequent resistance phenotypes throughout the study period but decreased from 27.8% in Year 1 to 20.9% in Year 4. Ampicillin resistance also declined, from 7.8% to 2.6%.

**Figure 4.** Temporal trends in antimicrobial resistance among pneumococcal isolates, 2022–2026 Lines represent annual resistance percentages for the indicated antimicrobial agents and multidrug resistance (MDR). Annual isolate denominators were 425, 376, 209, and 302 in Years 1–4, respectively, subject to the availability of valid susceptibility results for each agent. Both typeable and non-typeable isolates were included. Temporal trends were assessed using the Cochran–Armitage test for trend. Tests were exploratory because repeated isolates from individual children could contribute correlated observations. TMP-SMX, trimethoprim–sulfamethoxazole; MDR, multidrug resistance.

**Table 12.** Temporal trends in antimicrobial resistance among Streptococcus pneumoniae isolates across four surveillance years, Novi Sad, Serbia, 2022–2026

| Antimicrobial agent | Year 1 n/N ()<br>N=425 | Year 2 n/N ()<br>N=376 | Year 3 n/N ()<br>N=209 | Year 4 n/N (%)<br>N=302 | p for trend |
|---|---|---|---|---|---|
| Penicillin G | 1 (0.2) | 1 (0.3) | 9 (4.3) | 6 (2.0) | 0.001 |
| Ampicillin | 33 (7.8) | 20 (5.3) | 12 (5.7) | 8 (2.6) | 0.005 |
| Erythromycin | 128 (30.1) | 103 (27.4) | 62 (29.7) | 63 (20.9) | 0.014 |
| Clindamycin | 103 (24.2) | 84 (22.3) | 32 (15.3) | 42 (13.9) | <0.001 |
| Trimethoprim-sulfamethoxazole | 43 (10.1) | 15 (4.0) | 11 (5.3) | 67 (22.2) | <0.001 |
| Tetracycline | 92 (21.6) | 82 (21.8) | 30 (14.4) | 34 (11.3) | <0.001 |
| MDR | 96 (22.6) | 77 (20.5) | 30 (14.4) | 35 (11.6) | <0.001 |

Values are presented as the count and percentage of resistant pneumococcal isolates within each surveillance year. Annual denominators were 425, 376, 209, and 302 isolates in Years 1–4, respectively, including both typeable and non-typeable isolates. These denominators apply where susceptibility results were available for all isolates; otherwise, agent-specific denominators must be reported. Temporal trends were assessed using the Cochran–Armitage test for trend with ordered surveillance-year scores of 1–4. Repeated isolates from the same child were retained, and within-child correlation was not accounted for; therefore, inferential results were exploratory. Penicillin G results refer to non-susceptibility, not resistance. MDR was defined as resistance to at least one antimicrobial agent in three or more distinct antimicrobial classes.

In contrast, trimethoprim–sulfamethoxazole resistance increased markedly in Year 4, rising to 22.2% after lower levels in Years 2 and 3. Penicillin G non-susceptibility remained uncommon overall but increased after the first two years, reaching 4.3% in Year 3 and 2.0% in Year 4. These temporal patterns should be interpreted cautiously because repeated isolates from the same child were retained and within-child correlation was not accounted for in the trend analyses.

Analyses included both typeable and non-typeable isolates.

### Serotype-Specific Antimicrobial Resistance

Antimicrobial resistance varied substantially across pneumococcal serotype categories (Figure 5 and Supplementary Table S9). The highest erythromycin resistance proportions were observed among serotypes 6C (83.3%), 6B (81.0%), and 24F (75.0%); these serotypes also showed high levels of clindamycin and tetracycline resistance. Multidrug resistance was most frequent in serotypes 6C (66.7%), 24F (64.6%), and 6B (63.5%), indicating that resistance was concentrated in selected serotype groups. In absolute numbers, however, the largest contributions to MDR were from non-typeable isolates (n = 47), followed by serotypes 6B (n = 40), 24F (n = 31), 6C (n = 28), and 15A (n = 27).

**Figure 5.** Heatmap of antimicrobial resistance profiles by pneumococcal serotype among nasopharyngeal Streptococcus pneumoniae isolates in children, Novi Sad, Serbia, 2022–2026 Each cell represents the share of isolates resistant to the indicated antimicrobial agent or classified as multidrug-resistant (MDR) within the corresponding serotype category. Category denominators are displayed in parentheses and correspond to Table 19. The analysis includes the 15 most frequent typeable serotypes, other typeable serotypes grouped as “Others,” and non-typeable isolates. Percentages were computed using isolates with valid susceptibility results. The color gradient represents increasing resistance frequency. Results are descriptive and were not adjusted for repeated isolates from the same child.

Several serotypes, including 10A, 11A, 23A, 15B, 3, and 35F, showed comparatively low resistance frequencies across most antimicrobial agents. Serotype 23B generally showed low resistance to macrolides, clindamycin, tetracycline, and MDR, but had a relatively higher trimethoprim–sulfamethoxazole resistance proportion. Figure 5 is descriptive, and percentages were calculated among isolates with valid susceptibility results. Complete serotype-specific resistance data are presented in Supplementary Table S9.

### Vaccine-Serotype Distribution According to Vaccination Status

The distribution of pneumococcal isolates corresponding to licensed vaccine formulations was examined according to vaccination status (Supplementary Table S10).

PCV20-type serotypes accounted for 48.1% of typeable isolates from fully vaccinated children, 47.0% from partially vaccinated children, and 35.7% from unvaccinated children. Correspondingly, serotypes not included in PCV20 represented 51.9%, 53.0%, and 64.3% of isolates in these groups.

These descriptive findings were not interpreted as estimates of vaccine effectiveness.

## Discussion

This completed four-year surveillance study describes pneumococcal nasopharyngeal carriage among children aged 2– 60 months in Novi Sad from October 2022 through September 2026. Three findings were most prominent. First, carriage was common and was more frequent among children presenting with acute respiratory tract infection than among healthy children. Second, prevalence varied by age, season, and surveillance year, with the highest carriage prevalence observed among children aged 24–35 months and the lowest seasonal prevalence during summer. Third, more than half of typeable carriage isolates belonged to serotypes not included in PCV20, while resistance to erythromycin, clindamycin, and tetracycline remained frequent. Overall carriage prevalence was 27.0%, confirming that young children remain an important reservoir of S. pneumoniae in the community. Nasopharyngeal colonization is a prerequisite for pneumococcal transmission and generally precedes pneumococcal disease, but carriage is frequently asymptomatic and should not be considered equivalent to clinical infection (1,2).

The observed prevalence was lower than the 31.7% reported in a previous Novi Sad pilot study conducted during 2020– 2021 (46). However, direct comparison requires caution because the studies differed in age composition and epidemiological context. The earlier investigation included children aged 24–60 months, whereas the present study included children aged 2–60 months. Carriage prevalence was lowest among infants aged 2–11 months in the current surveillance, and their inclusion contributed to the lower overall estimate. In the present study, carriage prevalence in the age group 24–60 was 30.0% when the 24–35, 36–47, and 48–60 month age groups in Table 2 were combined.

International studies have demonstrated substantial variation in pneumococcal carriage prevalence according to geographic setting, age, vaccination history, and study design. In a systematic review including 82 studies, Neal et al. reported median carriage prevalence in the general population ranging from 28.5% in high-income countries to 51.0% in low-income countries. The prevalence observed in Novi Sad was therefore close to the median reported for high-income settings, although differences in sampling methods and study populations limit direct comparisons (74).

The higher carriage prevalence among children with acute respiratory tract infection is consistent with the role of pneumococci as common colonizers of the upper respiratory tract during early childhood. Respiratory viral infections may alter the nasopharyngeal environment, mucosal immune responses, and interactions between pneumococci and other respiratory microorganisms, potentially influencing bacterial acquisition, density, or persistence (74–78). Previous studies have demonstrated complex interactions between respiratory viruses and pneumococcal colonization, supporting the biological plausibility of increased pneumococcal detection during respiratory illness (74). Because nasopharyngeal detection reflects colonization rather than disease causation, these findings should be interpreted as differences in carriage between clinical groups, not as evidence that pneumococci caused the respiratory illness. Despite differences in carriage prevalence, the overall serotype distributions among isolates recovered from healthy and sick children were broadly similar. The weak association between clinical status and serotype composition suggests that the principal circulating serotypes were shared between the two groups (79). Individual-serotype comparisons were exploratory and should be interpreted cautiously, particularly when significant only before adjustment for multiple comparisons (80,81).

Age was strongly associated with carriage, with the highest prevalence observed among children aged 24–35 months. This pattern is biologically plausible because children in this age range have frequent close contact with peers, developing mucosal immunity, and high exposure to respiratory pathogens (74,82–85). Daycare attendance may contribute to this pattern, as previous investigations have identified childcare attendance and contact with other young children as factors associated with pneumococcal carriage (74,83–85). However, age and daycare exposure are closely related and may jointly contribute to the observed patterns (2,83,85). Because the analysis was primarily descriptive, the independent contribution of daycare attendance could not be fully separated from age. Chronic medical conditions were associated with higher carriage prevalence overall, but this finding was descriptive and based on a relatively small subgroup.

Carriage also varied by season. Prevalence was highest in autumn and winter, remained relatively high in spring, and was lowest in summer. This pattern is compatible with seasonal differences in indoor crowding, respiratory-virus circulation, and transmission opportunities (77,86–89). The summer decline was substantial, but the study was not designed to identify the specific mechanisms responsible for seasonal variation. The analysis of 16 consecutive seasons demonstrated that seasonal fluctuations were recurrent but not uniform across surveillance years. The highest overall seasonal prevalence was observed in autumn 2023, whereas the lowest occurred in summer 2024. Monthly analyses provided additional descriptive evidence of substantial short-term variation. These findings emphasize the importance of considering both calendar season and surveillance year when comparing carriage prevalence estimates, because differences in the seasonal composition of recruited children may influence annual estimates and complicate comparisons between surveillance periods (86,89).

Annual prevalence varied during the surveillance period, with lower prevalence in Year 3 and a partial increase in Year 4 rather than a sustained monotonic decline. Because the study overlapped with changes in pneumococcal vaccination policy, these results are useful for describing carriage patterns during a period of changing vaccine use. PCV13 replaced PCV10 for routine childhood immunization in April 2022, the vaccination schedule changed from 3+1 to 2+1 in January 2023, and PCV10 was reintroduced in January 2024 (44,45). These programmatic changes occurred in the context of evolving pneumococcal serotype distribution after childhood PCV introduction in Serbia (90). However, the study design does not allow the observed temporal changes to be attributed directly to vaccination-policy changes, secular variation, respiratory-virus activity, antibiotic use, or other population-level factors (91).

The pneumococcal isolate population exhibited considerable serotype diversity. Serotypes 11A, 10A, 15A, 15B, 23A, and 23B were among the most frequently recovered, while non-typeable isolates accounted for 12.1% of all isolates. The predominance of these serotypes is consistent with changing pneumococcal ecology reported in European countries following widespread pneumococcal conjugate vaccine use. Candeias et al. identified serotypes 15B/C, 11A, 23B, and 23A among the most prevalent non-PCV13 pneumococci in Portuguese children, while Horváth et al. reported frequent carriage of serotypes 23B, 35F, 15A/F, 15B/C, 11A, and 23A among children in Hungary. Both studies report serotype distributions in daycare attendees. Therefore, direct comparison is somewhat limited because our study did not investigate daycare attendees only, and provides broader and combined data analysis with those not attending daycare as well (39,92). However, the substantial representation of serotypes not included in earlier pneumococcal conjugate vaccines is epidemiologically important. In vaccinated populations, reductions in vaccine-included serotypes may alter competition within the nasopharyngeal ecological niche, allowing non-vaccine serotypes to become more prominent. This phenomenon is commonly described as serotype replacement (3,14,86,93).

In the present study, theoretical serotype coverage in licensed PCV formulations varied across surveillance years, while the relative frequencies of individual serotypes changed. In particular, serotype 23A showed a decreasing temporal trend, whereas serotype 23B increased. These findings are compatible with changes in pneumococcal population composition that may occur during serotype replacement (86,94). Serotypes also differ in their capacity for colonization, transmission, and invasive disease. Balsells et al. demonstrated substantial variation in serotype-specific invasive disease potential, indicating that the frequency of a serotype in nasopharyngeal carriage cannot be directly translated into its contribution to severe pneumococcal disease (95). Continued circulation of vaccine-included serotypes may reflect differences in serotype-specific vaccine effects, continued transmission, or coexistence of children from different vaccination cohorts (14,96).

The serotype distribution was dominated by non-PCV20 serotypes among typeable carriage isolates. Among typeable isolates, 8.2% corresponded to PCV10 serotypes, 17.5% to PCV13, 20.0% to PCV15, and 47.3% to PCV20. Nevertheless, 52.7% of typeable isolates belonged to serotypes not included in PCV20, demonstrating substantial pneumococcal diversity beyond the serotype composition of the evaluated licensed formulations (94,97). The theoretical serotype coverage of circulating isolates in PCV20 was 47.3% in Novi Sad, compared with 30.5% reported by Candeias et al. in Portugal and 31.8% reported by Horváth et al. in Hungary (39,92). These differences require cautious interpretation because the studies differed in surveillance periods, age groups, recruitment settings, and vaccination histories. As expected, higher-valency formulations included a larger proportion of observed carriage serotypes than lower-valency formulations. Based on the formulations evaluated in Supplementary Table S1, the highest theoretical serotype coverage was observed for Pn-MAPS30plus, 25vPnC/PF-07872412, and VAX-31, and Pn-MAPS30plus covered the greatest proportion of serotypes not covered by PCV20. However, theoretical serotype coverage should be interpreted only as serotype match. It does not account for vaccine uptake, indirect effects, invasiveness of individual serotypes, immune response, or clinical outcomes. These findings therefore describe potential coverage of carried serotypes, not expected vaccine impact (3,14,27,82,95,98).

Antimicrobial resistance was common among carried pneumococci, particularly to erythromycin, clindamycin, and tetracycline, followed by trimethoprim–sulfamethoxazole. Penicillin G non-susceptibility was uncommon, and no levofloxacin resistance was detected. Similar resistance patterns have been reported in previous pneumococcal carriage investigations (99,100). The predominance of resistance to macrolides, lincosamides, and tetracyclines was broadly consistent with the previous Novi Sad pilot study, in which resistance to erythromycin, clindamycin, and tetracycline exceeded 30% (46). The lower resistance proportions observed in the present surveillance may reflect differences in circulating serotypes or resistance characteristics, although direct comparisons are limited by differences in age composition and surveillance periods.

Multidrug resistance was observed in a substantial minority of circulating pneumococci, indicating resistance to antimicrobial agents belonging to at least three classes. Resistant pneumococci carried in the nasopharynx may contribute to the community reservoir of antimicrobial resistance without causing immediate disease (1,101). In the comparison between PCV20 vaccine-type and non-vaccine-type isolates, ampicillin resistance was significantly higher among vaccine-type isolates, whereas the remaining predefined resistance outcomes did not differ significantly after adjustment for multiple comparisons. Temporal resistance patterns were heterogeneous, with lower proportions of resistance to several antimicrobial agents and multidrug resistance observed during later surveillance periods, whereas trimethoprim–sulfamethoxazole resistance increased toward the final surveillance year. Because these analyses included repeated isolates from some children, temporal patterns should be interpreted as exploratory changes in the recovered isolate population rather than independent longitudinal changes in individual children.

Resistance patterns also varied across pneumococcal serotypes. The highest observed proportions of multidrug resistance occurred among serotypes 6C, 24F, and 6B, with relatively elevated proportions also recorded among serotypes 19A and 15A and non-typeable isolates. However, these descriptive differences should be interpreted cautiously, particularly for less frequent serotypes. In absolute terms, non-typeable isolates contributed the largest number of multidrug-resistant isolates, followed by serotypes 6B, 24F, 6C, and 15A. Conversely, several frequently circulating serotypes, including 10A, 11A, 23A, 15B, 3, and 35F, generally exhibited low resistance frequencies across the antimicrobial agents tested. Serotype 23B showed comparatively low resistance to macrolides, clindamycin, tetracycline, and multidrug resistance, but a relatively higher proportion of trimethoprim–sulfamethoxazole resistance. These findings suggest that serotype prevalence alone may not adequately reflect the antimicrobial resistance burden, as less prevalent serotypes may exhibit relatively high resistance proportions while more prevalent serotypes may remain largely susceptible (32,102,103). Previous studies have demonstrated associations between pneumococcal serotypes, genetic lineages, and antimicrobial resistance, including the persistence or emergence of resistant lineages among non- vaccine serotypes following conjugate vaccine introduction (32,34,98,101,103,104). However, the genetic mechanisms underlying the observed resistance patterns could not be assessed because molecular characterization of pneumococcal lineages was not performed. Similarly, the potential contribution of antimicrobial selective pressures could not be evaluated without antimicrobial consumption data. These findings support continued surveillance of pneumococcal antimicrobial susceptibility, ideally complemented by molecular characterization and information on antimicrobial use. Vaccination-status comparisons were descriptive. Overall pneumococcal carriage prevalence did not differ significantly according to recorded vaccination status, and neither the overall comparison nor the ordered trend was statistically significant. This finding should not be interpreted as evidence that pneumococcal vaccination has no effect on carriage, because vaccination status may be associated with age, birth cohort, vaccine formulation, time since vaccination, healthcare-seeking behavior, and other factors that were not fully adjusted for (86,103). Previous studies have shown that pneumococcal conjugate vaccines reduce carriage of vaccine-included serotypes, while increases in non-vaccine-type carriage may attenuate changes in overall carriage prevalence (14,26,27,103). In the isolate-level analysis, PCV20- type serotypes represented 48.1% of typeable isolates from fully vaccinated children, 47.0% from partially vaccinated children, and 35.7% from unvaccinated children. These proportions describe the composition of recovered isolates within vaccination categories, not vaccine effectiveness or vaccination-specific carriage risk. The unvaccinated category contributed a comparatively small number of typeable isolates, and repeated isolates were retained without adjustment for within-child correlation (105).

The main strengths of this study are its prospective design, completed four-year surveillance period, inclusion of both healthy children and children with acute respiratory tract infection, and combined analysis of carriage prevalence, serotype distribution, and antimicrobial susceptibility. These features provide a detailed local description of pneumococcal carriage among young children in Novi Sad.

This study also has limitations. Recruitment was consecutive within predefined age strata and was conducted in one city, which may limit generalizability to other regions of Serbia. Because several potentially important confounders, including number of older siblings and household smoking exposure, were not collected, the analysis remained primarily descriptive and multivariable models of independent carriage predictors were not fitted. Participant-level prevalence analyses used the first eligible specimen from each child, whereas isolate-level analyses included isolates from repeated visits and were therefore considered exploratory. The study was not designed to estimate vaccine effectiveness, and vaccination-status comparisons were not adjusted for all potential confounders. Only one colony was selected per culture, so co-colonization with multiple pneumococcal serotypes may have been missed (106). In addition, children who had received systemic antibiotics within seven days before sampling were excluded, which may have influenced prevalence and resistance estimates (100,107). Pneumococcal density, respiratory viral coinfections in sick children, detailed antibiotic-use data, and molecular characteristics of resistant isolates were not systematically investigated.

Finally, carriage does not necessarily indicate disease causation, and carriage serotype distributions or theoretical vaccine coverage should not be interpreted as direct estimates of invasive disease burden or vaccine impact (88,95,98).

## Conclusions

In this completed four-year surveillance study of children aged 2–60 months in Novi Sad, pneumococcal carriage was associated with age, clinical presentation, season, and surveillance year. More than half of typeable carriage isolates were non-PCV20 serotypes, and resistance to erythromycin, clindamycin, and tetracycline was common. These findings provide local evidence on pneumococcal carriage and resistance patterns during a period of changing vaccine use.

Continued carriage surveillance, linked where possible with invasive-disease and antibiotic-use data, would help determine whether the observed serotype and resistance patterns persist.

## Supporting information

Supplementary Figure S1. Temporal trend in carriage prevalence by surveillance year, season and clinical status

Supplementary Figure S2.Temporal trend in carriage prevalence by surveillance months and clinical status

Supplementary Table S1. Serotype inclusion in licensed and investigational pneumococcal conjugate vaccines

Supplementary Table S2. Pneumococcal carriage prevalence in healthy versus sick children by demographic and clinical characteristics

Supplementary Table S3. Distribution of Streptococcus pneumoniae serotypes among healthy and sick children and serotype-specific associations with hea

Supplementary Table S4. Participant-level pneumococcal carriage prevalence by surveillance year

Supplementary Table S5. Participant-level pneumococcal carriage prevalence by surveillance year and clinical status

Supplementary Table S6. Serotype inclusion of pneumococcal isolates in licensed pneumococcal conjugate vaccine formulations by surveillance years

Supplementary Table S7. Antimicrobial resistance by clinical presentation

Supplementary Table S8. Antimicrobial resistance in VT and NVT isolates: unadjusted and Holm-adjusted p-values

Supplementary Table S9. Distribution of antimicrobial resistance and multidrug resistance among Streptococcus pneumoniae serotypes isolated from child

## Data Availability

All data produced in the present study are available upon reasonable request to the authors

## Acknowledgments

We thank the laboratory technicians at the Institute of Public Health of Vojvodina, Novi Sad who participated in this project.

## Supporting infomation

Supplementary Table S2. Pneumococcal carriage prevalence in healthy versus sick children by demographic and clinical characteristics Carriage prevalence was calculated as the count of culture-positive children divided by the count of children tested within each demographic or clinical category. Each child contributed the first eligible nasopharyngeal specimen. Ninety-five percent confidence intervals were estimated using the Clopper–Pearson exact binomial method P-values compare carriage prevalence between healthy and sick children within each stratum and were calculated using Pearson’s chi-square test or Fisher’s exact test, as appropriate. CI, confidence interval.

Supplementary Table S3. Distribution of Streptococcus pneumoniae serotypes among healthy and sick children and serotype-specific associations with health status Overall serotype distribution: Pearson χ²(47) = 50.02, p = 0.354, Cramér’s V = 0.195. ¹ OR compares the odds of each serotype versus all other serotypes among healthy relative to sick children. Thus, OR >1 indicates greater relative representation in healthy children, whereas OR <1 indicates greater relative representation in sick children. ² Two-sided Fisher’s exact test. ³ Benjamini–Hochberg correction was applied across the 48 serotype-specific tests to control the false discovery rate (FDR). The BH procedure adjusts the set of individual p-values for multiple testing ⁴ Because one cell contained zero isolates, the conventional OR is 0 or infinity and is not meaningfully estimable with ordinary asymptotic methods. NT, non-typeable.

Supplementary Figure S1. Temporal trend in carriage prevalence by surveillance year, season and clinical status Bars represent the seasonal prevalence (%) of nasopharyngeal Streptococcus pneumoniae carriage among all children (grey), healthy children (blue), and sick children (red). Error bars indicate 95% confidence intervals (CIs) calculated using the Wilson score method without continuity correction. Seasons were defined as autumn (October–December), winter (January–March), spring (April–June), and summer (July–September). Vertical dashed lines separate the four surveillance years, each extending from October through September, from 2022 to 2026. Carriage prevalence was calculated as the proportion of children with culture-confirmed S. pneumoniae carriage among all children tested in each season and clinical group.

Supplementary Figure S2. Temporal trend in carriage prevalence by surveillance months and clinical status Bars represent the monthly prevalence (%) of nasopharyngeal Streptococcus pneumoniae carriage among all children (grey), healthy children (blue), and sick children (red). Error bars indicate 95% confidence intervals (CIs) calculated using the Wilson score method without continuity correction. Vertical dashed lines separate the four surveillance years (October–September), covering October 2022 to September 2026. Carriage prevalence was calculated as the proportion of children with culture-confirmed S. pneumoniae carriage among those tested in each month and clinical group.

Supplementary Table S4. Participant-level pneumococcal carriage prevalence by surveillance year Carriage prevalence was calculated using the first eligible specimen per unique child. Annual denominators were 1,233, 875, 643, and 622 children in Years 1–4, respectively. Ninety-five percent confidence intervals were estimated using the Clopper–Pearson exact binomial method. Overall differences across surveillance years were assessed using Pearson’s chi-square test, and ordered temporal trends using the Cochran–Armitage test for trend. Surveillance-year associations should not be taken as causal effects of changes in pneumococcal vaccination policy.

Supplementary Table S5. Participant-level pneumococcal carriage prevalence by surveillance year and clinical status Carriage prevalence was calculated using the first eligible specimen per unique child. Annual denominators were 1,233, 875, 643, and 622 children in Years 1–4, respectively. The corresponding annual denominators were 634, 449, 321, and 326 for healthy children and 599, 426, 322, and 296 for sick children. Ninety-five percent confidence intervals were estimated using the Clopper–Pearson exact binomial method. Overall differences across surveillance years were assessed using Pearson’s chi-square test, and ordered temporal trends using the Cochran–Armitage test for trend. Annual p-values compare healthy and sick children within the same surveillance year, while overall and trend p-values compare surveillance years within each clinical-status group. Surveillance-year associations should not be taken as causal effects of changes in pneumococcal vaccination policy.

Supplementary Table S6. Serotype inclusion of pneumococcal isolates in licensed pneumococcal conjugate vaccine formulations by surveillance years Values are presented as numbers (n), percentages (%), and 95% confidence intervals (CIs). Percentages were computed using the total number of typeable pneumococcal isolates recovered during each surveillance year. The 95% CIs were estimated using the Clopper–Pearson exact binomial method. Differences in vaccine-type isolate proportions across surveillance years were assessed using Pearson’s chi-square test, while linear temporal trends were assessed using the Cochran–Armitage test for trend, Vaccine-type categories represent cumulative serotype inclusion in the respective PCV formulations and are not mutually exclusive. Repeated isolates from the same child were included; therefore, inferential tests were exploratory and did not account for within-child correlation.

Supplementary Table S7. Antimicrobial resistance by clinical presentation Values are presented as n (%) of isolates resistant to each antimicrobial agent within the corresponding clinical-status group. The isolate-level dataset included 536 isolates from healthy children and 776 isolates from sick children, including typeable and non-typeable isolates. For each agent, the denominator should correspond to the count of isolates with a valid susceptibility result. Comparisons were performed using Pearson’s chi-square test or Fisher’s exact test, as appropriate. Reported p-values are unadjusted unless otherwise specified. Repeated isolates from the same child could contribute correlated observations; therefore, comparisons were exploratory. MDR, multidrug resistance.

Supplementary Table S8. Antimicrobial resistance in VT and NVT isolates: unadjusted and Holm-adjusted p-values Data are presented as the number and percentage [n (%)] of resistant Streptococcus pneumoniae isolates. Percentages were computed using the total number of PCV20 vaccine-type (VT; n = 545) or non-vaccine-type (NVT; n = 608) isolates as the denominator. Non-typeable isolates (n = 159) were excluded from the analysis. Unadjusted p-values were computed using Pearson’s chi-square test without continuity correction, except for penicillin G, for which Fisher’s exact test was applied. P-values were adjusted using the Holm–Bonferroni step-down procedure to control the family-wise error rate across seven predefined comparisons (six antimicrobial agents and multidrug resistance [MDR]). Statistical significance was defined as a Holm-adjusted p-value < 0.05.MDR was defined as resistance to at least one antimicrobial agent in three or more distinct antimicrobial classes. Oxacillin screening positivity alone was not considered confirmed β-lactam resistance for MDR classification. Penicillin G results refer to non- susceptibility, not resistance. Because repeated isolates from the same child could be included, isolate-level comparisons were regarded as exploratory and did not account for within-child correlation. Abbreviations: PCV20, 20-valent pneumococcal conjugate vaccine; VT, vaccine-type; NVT, non-vaccine-type; MDR, multidrug resistance.

Supplementary Table S9. Distribution of antimicrobial resistance and multidrug resistance among Streptococcus pneumoniae serotypes isolated from children in Novi Sad, Serbia, 2022–2026. Values are presented as the count and percentage of resistant isolates within each serotype category. The table includes 1,312 pneumococcal isolates, comprising 1,153 typeable isolates and 159 non-typeable isolates. The 15 most frequent typeable serotypes are presented individually, while remaining typeable serotypes are grouped as “Others.” Non-typeable isolates are presented separately. For each antimicrobial agent, the percentage denominator was the count of isolates within the corresponding serotype category with a valid susceptibility result. MDR was defined as resistance to at least one antimicrobial agent in three or more distinct antimicrobial classes. Resistance was interpreted using applicable EUCAST clinical breakpoints. Analyses were descriptive, and repeated isolates from the same child were retained.

Supplementary Table S10. Distribution of pneumococcal vaccine-type isolates according to vaccination status among children aged 2–60 months, Novi Sad, Serbia, 2022–2026. Data are presented as numbers and percentages of pneumococcal isolates within each vaccination category. Repeated isolates from the same child were included; therefore, observations could not be assumed to be independent. The analysis was descriptive, and no statistical significance testing was performed, Vaccine-type categories represent cumulative serotype coverage of the respective PCV formulations, PCV20-NVT denotes isolates not belonging to PCV20 vaccine serotypes.

