## Supplementary figures and images for "Pneumococcal nasopharyngeal carriage in children 2–60 months of age in Novi Sad (Serbia), October 2022–September 2026"

### Supplementary Figure S1. Temporal trend in carriage prevalence by surveillance year, season and clinical status

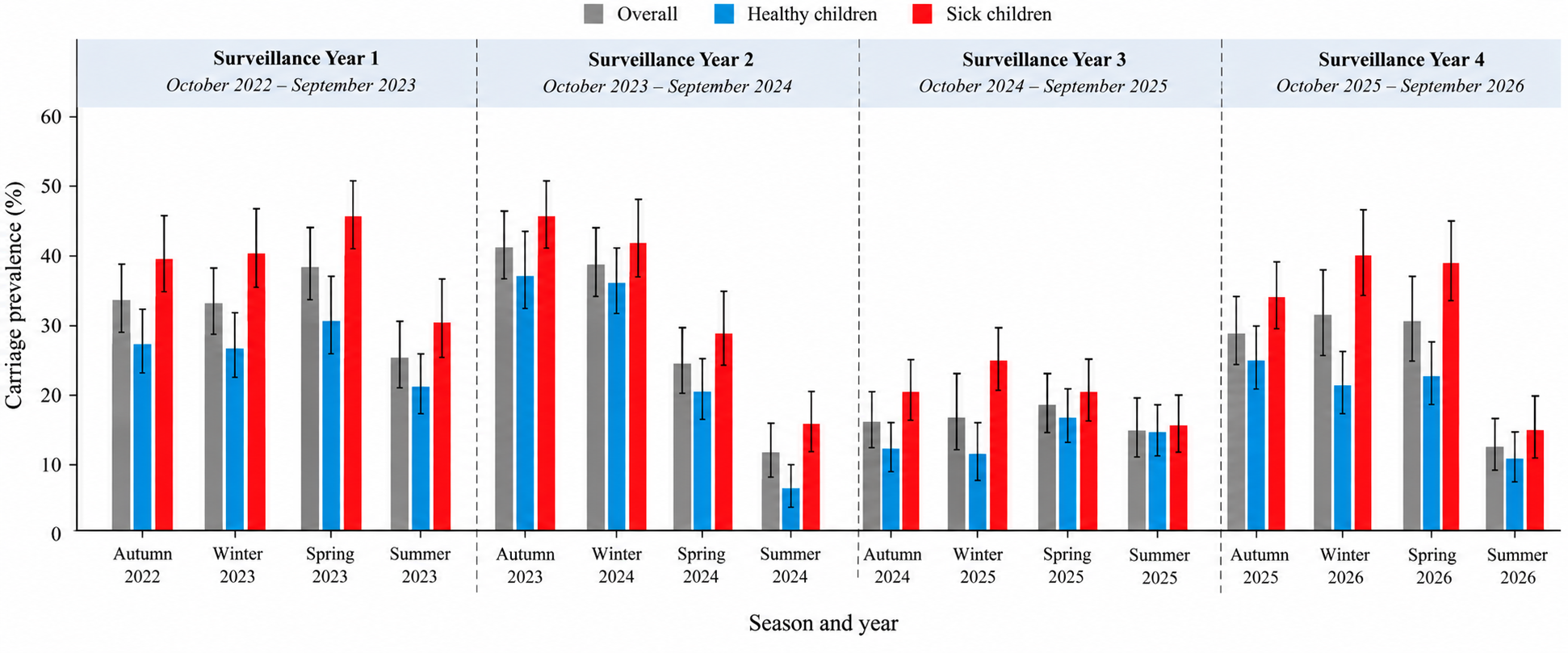

### Supplementary Figure S2.Temporal trend in carriage prevalence by surveillance months and clinical status

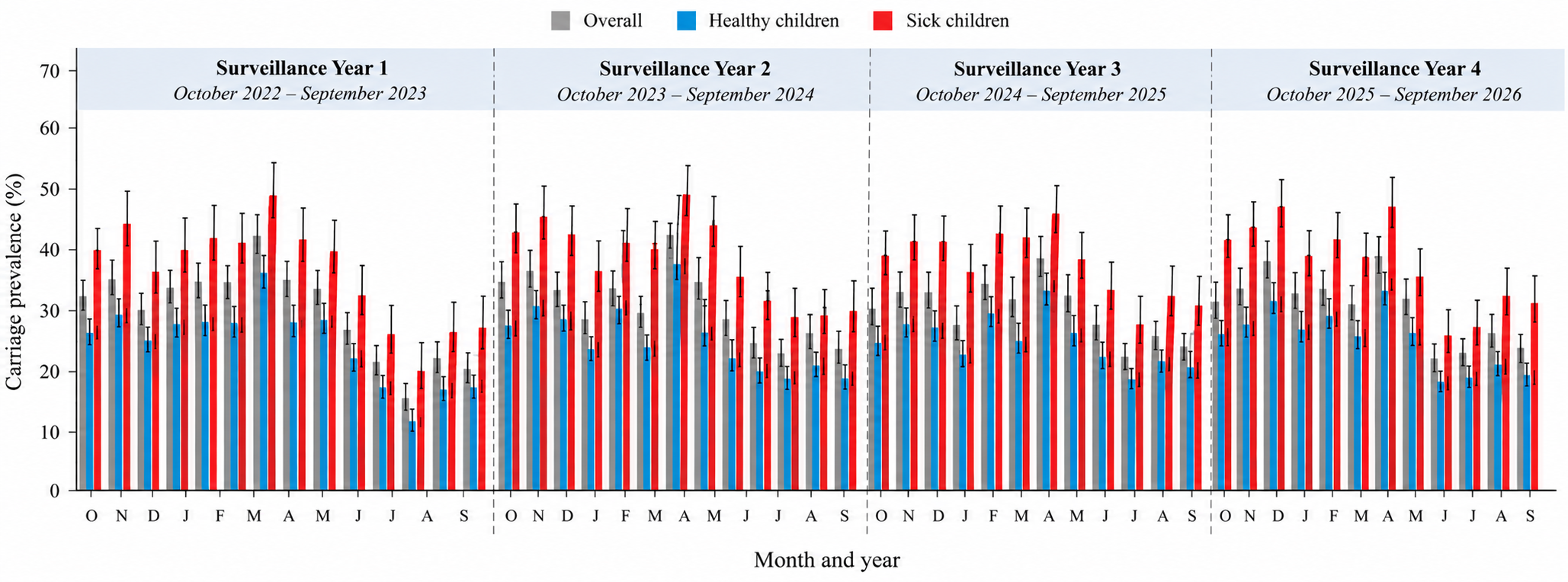
