## Supplementary Table S2. Pneumococcal carriage prevalence in healthy versus sick children by demographic and clinical characteristics for "Pneumococcal nasopharyngeal carriage in children 2–60 months of age in Novi Sad (Serbia), October 2022–September 2026"

| **Characteristic** | **Category** | **Healthy, positive/tested (%)** | **95% CI** | **Sick, positive/tested (%)** | **95% CI** | **p-value for healthy vs. sick comparison** |
| --- | --- | --- | --- | --- | --- | --- |
| Overall | — | 381/1730 (22.0) | 20.0–24.0 | 530/1643 (32.3) | 30.0–34.6 | <0.001 |
| Age group | 2–11 months | 79/521 (15.2) | 12.1–18.2 | 134/466 (28.8) | 24.6–32.9 | <0.001 |
|  | 12–23 months | 72/367 (19.6) | 15.6–23.7 | 131/370 (35.4) | 30.5–40.3 | <0.001 |
|  | 24–35 months | 78/274 (28.5) | 23.1–33.8 | 101/284 (35.6) | 30.0–41.1 | 0.072 |
|  | 36–47 months | 76/287 (26.5) | 21.4–31.6 | 79/243 (32.5) | 26.6–38.4 | 0.129 |
|  | 48–60 months | 76/281 (27.0) | 21.9–32.2 | 85/280 (30.4) | 25.0–35.7 | 0.386 |
| Sex | Male | 193/908 (21.3) | 18.6–23.9 | 276/888 (31.1) | 28.0–34.1 | <0.001 |
|  | Female | 188/822 (22.9) | 20.0–25.7 | 254/755 (33.6) | 30.3–37.0 | <0.001 |
| Daycare attendance | Yes | 266/932 (28.5) | 25.6–31.4 | 343/980 (35.0) | 32.0–38.0 | 0.002 |
|  | No | 115/798 (14.4) | 12.0–16.8 | 187/663 (28.2) | 24.8–31.6 | <0.001 |
| Chronic medical condition | Yes | 19/85 (22.4) | 13.5–31.2 | 46/108 (42.6) | 33.3–51.9 | 0.003 |
|  | No | 362/1645 (22.0) | 20.0–24.0 | 484/1535 (31.5) | 29.2–33.9 | <0.001 |
| Vaccination status | Fully vaccinated | 225/1067 (21.1) | 18.6–23.5 | 358/1143 (31.3) | 28.6–34.0 | <0.001 |
|  | Partially vaccinated | 133/545 (24.4) | 20.8–28.0 | 151/423 (35.7) | 31.1–40.3 | <0.001 |
|  | Unvaccinated | 23/118 (19.5) | 12.3–26.6 | 21/77 (27.3) | 17.3–37.2 | 0.203 |
