## Supplementary Table S3. Distribution of Streptococcus pneumoniae serotypes among healthy and sick children and serotype-specific associations with hea for "Pneumococcal nasopharyngeal carriage in children 2–60 months of age in Novi Sad (Serbia), October 2022–September 2026"

| **Serotype** | **Overall n (%)** | **Healthy n (%)** | **Sick n (%)** | **OR (95% CI)¹** | **p value²** | **FDR p³** |
| --- | --- | --- | --- | --- | --- | --- |
| NT | 159 (12.1) | 62 (11.6) | 97 (12.5) | 0.92 (0.65–1.29) | 0.667 | 1.000 |
| 11A | 111 (8.5) | 46 (8.6) | 65 (8.4) | 1.03 (0.69–1.52) | 0.920 | 1.000 |
| 10A | 94 (7.2) | 38 (7.1) | 56 (7.2) | 0.98 (0.64–1.50) | 1.000 | 1.000 |
| 15A | 93 (7.1) | 42 (7.8) | 51 (6.6) | 1.21 (0.79–1.85) | 0.384 | 1.000 |
| **15B** | **82 (6.3)** | **24 (4.5)** | **58 (7.5)** | **0.58 (0.36–0.95)** | **0.028** | **1.000** |
| 23A | 82 (6.3) | 38 (7.1) | 44 (5.7) | 1.27 (0.81–1.99) | 0.299 | 1.000 |
| 23B | 81 (6.2) | 35 (6.5) | 46 (5.9) | 1.11 (0.70–1.75) | 0.727 | 1.000 |
| 19A | 64 (4.9) | 26 (4.9) | 38 (4.9) | 0.99 (0.59–1.65) | 1.000 | 1.000 |
| 6B | 63 (4.8) | 30 (5.6) | 33 (4.3) | 1.33 (0.80–2.22) | 0.294 | 1.000 |
| 15C | 52 (4.0) | 19 (3.5) | 33 (4.3) | 0.83 (0.47–1.47) | 0.567 | 1.000 |
| 21 | 51 (3.9) | 20 (3.7) | 31 (4.0) | 0.93 (0.53–1.65) | 0.885 | 1.000 |
| 24F | 48 (3.7) | 20 (3.7) | 28 (3.6) | 1.04 (0.58–1.86) | 1.000 | 1.000 |
| 35B | 42 (3.2) | 19 (3.5) | 23 (3.0) | 1.20 (0.65–2.23) | 0.633 | 1.000 |
| 6C | 42 (3.2) | 21 (3.9) | 21 (2.7) | 1.47 (0.79–2.71) | 0.264 | 1.000 |
| 3 | 32 (2.4) | 12 (2.2) | 20 (2.6) | 0.87 (0.42–1.79) | 0.856 | 1.000 |
| 35F | 29 (2.2) | 12 (2.2) | 17 (2.2) | 1.02 (0.48–2.16) | 1.000 | 1.000 |
| 8 | 23 (1.8) | 12 (2.2) | 11 (1.4) | 1.59 (0.70–3.64) | 0.289 | 1.000 |
| 22F | 17 (1.3) | 3 (0.6) | 14 (1.8) | 0.31 (0.09–1.07) | 0.079 | 1.000 |
| 25A | 16 (1.2) | 4 (0.7) | 12 (1.5) | 0.48 (0.15–1.49) | 0.306 | 1.000 |
| 19F | 16 (1.2) | 8 (1.5) | 8 (1.0) | 1.45 (0.54–3.90) | 0.456 | 1.000 |
| 7C | 14 (1.1) | 9 (1.7) | 5 (0.6) | 2.63 (0.88–7.90) | 0.099 | 1.000 |
| 33F | 12 (0.9) | 4 (0.7) | 8 (1.0) | 0.72 (0.22–2.41) | 0.771 | 1.000 |
| 6A | 12 (0.9) | 4 (0.7) | 8 (1.0) | 0.72 (0.22–2.41) | 0.771 | 1.000 |
| 16F | 12 (0.9) | 6 (1.1) | 6 (0.8) | 1.45 (0.47–4.53) | 0.564 | 1.000 |
| 23F | 6 (0.5) | 0 (0.0) | 6 (0.8) | 0.00⁴ | 0.087 | 1.000 |
| 35A | 6 (0.5) | 4 (0.7) | 2 (0.3) | 2.91 (0.53–15.94) | 0.233 | 1.000 |
| 34 | 5 (0.4) | 2 (0.4) | 3 (0.4) | 0.97 (0.16–5.80) | 1.000 | 1.000 |
| 12F | 4 (0.3) | 0 (0.0) | 4 (0.5) | 0.00⁴ | 0.150 | 1.000 |
| 14 | 4 (0.3) | 1 (0.2) | 3 (0.4) | 0.48 (0.05–4.64) | 0.649 | 1.000 |
| 36 | 4 (0.3) | 1 (0.2) | 3 (0.4) | 0.48 (0.05–4.64) | 0.649 | 1.000 |
| 38 | 4 (0.3) | 1 (0.2) | 3 (0.4) | 0.48 (0.05–4.64) | 0.649 | 1.000 |
| 28 | 4 (0.3) | 3 (0.6) | 1 (0.1) | 4.36 (0.45–42.05) | 0.311 | 1.000 |
| 20 | 3 (0.2) | 0 (0.0) | 3 (0.4) | 0.00⁴ | 0.275 | 1.000 |
| 7F | 3 (0.2) | 1 (0.2) | 2 (0.3) | 0.72 (0.07–8.00) | 1.000 | 1.000 |
| 9L | 3 (0.2) | 1 (0.2) | 2 (0.3) | 0.72 (0.07–8.00) | 1.000 | 1.000 |
| 9N | 3 (0.2) | 1 (0.2) | 2 (0.3) | 0.72 (0.07–8.00) | 1.000 | 1.000 |
| 31 | 2 (0.2) | 0 (0.0) | 2 (0.3) | 0.00⁴ | 0.516 | 1.000 |
| 37 | 2 (0.2) | 0 (0.0) | 2 (0.3) | 0.00⁴ | 0.516 | 1.000 |
| 39 | 2 (0.2) | 1 (0.2) | 1 (0.1) | 1.45 (0.09–23.21) | 1.000 | 1.000 |
| 17F | 2 (0.2) | 2 (0.4) | 0 (0.0) | ∞⁴ | 0.167 | 1.000 |
| 13 | 1 (0.1) | 0 (0.0) | 1 (0.1) | 0.00⁴ | 1.000 | 1.000 |
| 41 | 1 (0.1) | 0 (0.0) | 1 (0.1) | 0.00⁴ | 1.000 | 1.000 |
| 42 | 1 (0.1) | 0 (0.0) | 1 (0.1) | 0.00⁴ | 1.000 | 1.000 |
| 9A | 1 (0.1) | 0 (0.0) | 1 (0.1) | 0.00⁴ | 1.000 | 1.000 |
| 1 | 1 (0.1) | 1 (0.2) | 0 (0.0) | ∞⁴ | 0.409 | 1.000 |
| 2 | 1 (0.1) | 1 (0.2) | 0 (0.0) | ∞⁴ | 0.409 | 1.000 |
| 40 | 1 (0.1) | 1 (0.2) | 0 (0.0) | ∞⁴ | 0.409 | 1.000 |
| 18C | 1 (0.1) | 1 (0.2) | 0 (0.0) | ∞⁴ | 0.409 | 1.000 |
| **Total** | **1.312 (100)** | **536 (100)** | **776 (100)** | — | — | — |
