## Supplementary Table S4. Participant-level pneumococcal carriage prevalence by surveillance year for "Pneumococcal nasopharyngeal carriage in children 2–60 months of age in Novi Sad (Serbia), October 2022–September 2026"

| Vaccine program period | Schedule | Surveillance year | Children tested, n | Positive, n | Prevalence % | 95% CI Lower limit | 95% CI Upper limit | P value | P for trend |
| --- | --- | --- | --- | --- | --- | --- | --- | --- | --- |
| PCV13 period | 3+1, 2+1 | Year 1: Oct 2022–Sep 2023 | 1233 | 396 | **32,1** | 29.5 | 34.7 | <0.001 | <0.001 |
| PCV13/PCV10 period | 2+1 | Year 2: Oct 2023–Sep 2024 | 875 | 255 | **29,1** | 26.1 | 32.2 |  |  |
| PCV10 period | 2+1 | Year 3: Oct 2024–Sep 2025 | 643 | 105 | **16,3** | 13.5 | 19.2 |  |  |
| PCV10 period | 2+1 | Year 4: Oct 2025–Sep 2026 | 622 | 155 | **24,9** | 21.5 | 28.3 |  |  |
