## Supplementary Table S5. Participant-level pneumococcal carriage prevalence by surveillance year and clinical status for "Pneumococcal nasopharyngeal carriage in children 2–60 months of age in Novi Sad (Serbia), October 2022–September 2026"

| Vaccine program period | Schedule | Surveillance year | Healthy | | | | | Sick | | | | | p value |
| --- | --- | --- | --- | --- | --- | --- | --- | --- | --- | --- | --- | --- | --- |
|  |  |  | Children tested, n | Positive, n | Prevalence % | 95% CI Lower limit | 95% CI Upper limit | Children tested, n | Positive, n | Prevalence % | 95% CI Lower limit | 95% CI Upper limit |  |
| PCV13 period | 3+1  2+1 | Year 1: Oct 2022–Sep 2023 | 634 | 164 | **25.9** | 22.5 | 29,3 | 599 | 232 | **38.7** | 34.8 | 42.6 | <0.001 |
| PCV13/PCV10 period | 2+1 | Year 2: Oct 2023–Sep 2024 | 449 | 113 | **25.2** | 21.2 | 29,2 | 426 | 142 | **33.3** | 28.9 | 37.8 | 0.010 |
| PCV10 period | 2+1 | Year 3: Oct 2024–Sep 2025 | 321 | 42 | **13.1** | 9.4 | 16,8 | 322 | 63 | **19.6** | 15.2 | 23.9 | 0.034 |
| PCV10 period | 2+1 | Year 4: Oct 2025–Sep 2026 | 326 | 62 | **19.0** | 14.8 | 23,3 | 296 | 93 | **31.4** | 26.1 | 36.7 | 0.001 |
| P value across surveillance years | | | <0.001 | | | | | <0.001 | | | | |  |
| p for trend | | | <0.001 | | | | | <0.001 | | | | |  |
