## Supplementary Table S6. Serotype inclusion of pneumococcal isolates in licensed pneumococcal conjugate vaccine formulations by surveillance years for "Pneumococcal nasopharyngeal carriage in children 2–60 months of age in Novi Sad (Serbia), October 2022–September 2026"

| Vaccine formulation | Year 1  n=363 | | Year 2  n=329 | | Year 3  N=187 | | Year 4  N=274 | | P value | P for trend |
| --- | --- | --- | --- | --- | --- | --- | --- | --- | --- | --- |
|  | Included serotype isolates, n (%) | 95% CI | Included serotype isolates, n (%) | 95% CI | Included serotype isolates, n (%) | 95% CI | Included serotype isolates, n (%) | 95% CI |  |  |
| PCV10 | 62 (17.1) | 13.4-21.4 | 23 (7.0) | 4.5-10.3 | 2 (1.1) | 0.1-3.8 | 7 (2.6) | 1.0-5.2 | <0.001 | <0.001 |
| PCV13 | 107 (29.5) | 24.8-34.5 | 46 (14.0) | 10.4-18.2 | 20 (10.7) | 6.7-16.0 | 29 (10.6) | 7.2-14.8 | <0.001 | <0.001 |
| PCV15 | 116 (32.0) | 27.2-37.0 | 50 (15.2) | 11.5-19.5 | 27 (14.4) | 9.7-20.3 | 38 (13.9) | 10.0-18.5 | <0.001 | <0.001 |
| PCV20 | 214 (59.0) | 53.7-64.1 | 145 (44.1) | 38.6-49.6 | 84 (44.9) | 37.7-52.3 | 102 (37.2) | 31.5-43.2 | <0.001 | <0.001 |
