## Supplementary Table S7. Antimicrobial resistance by clinical presentation for "Pneumococcal nasopharyngeal carriage in children 2–60 months of age in Novi Sad (Serbia), October 2022–September 2026"

| Antimicrobial agent | Total number of isolates in Healthy children, n | Healthy isolates resistant n (%) | Total number of isolates in Sick children, n | Sick isolates resistant n (%) | p-value |
| --- | --- | --- | --- | --- | --- |
| Penicillin G | 536 | 8 (1.5) | 776 | 9 (1.2) | 0.626 |
| Ampicillin | 536 | 25 (4.7) | 776 | 48 (6.2) | 0.237 |
| Erythromycin | 536 | 147 (27.4) | 776 | 209 (26.9) | 0.844 |
| Clindamycin | 536 | 116 (21.6) | 776 | 145 (18.7) | 0.187 |
| Trimethoprim-sulfamethoxazole | 536 | 58 (10.8) | 776 | 78 (10.1) | 0.653 |
| Tetracycline | 536 | 98 (18.3) | 776 | 140 (18.0) | 0.911 |
| MDR | 536 | 98 (18.3) | 776 | 140 (18.0) | 0.911 |
