## Supplementary Table S8. Antimicrobial resistance in VT and NVT isolates: unadjusted and Holm-adjusted p-values for "Pneumococcal nasopharyngeal carriage in children 2–60 months of age in Novi Sad (Serbia), October 2022–September 2026"

| Antimicrobial agent | VT isolates resistant n (%)  Total VT PCV20 isolates n=545 | NVT isolates resistant n (%)  Total NVT isolates n=608 | Unadjusted p | Holm-adjusted p |
| --- | --- | --- | --- | --- |
| Penicillin G | 4 (0.7) | 7 (1.2) | 0.554 | 1.000 |
| Ampicillin | 33 (6.1) | 13 (2.1) | <0.001 | 0.005 |
| Erythromycin | 122 (22.4) | 149 (24.5) | 0.396 | 1.000 |
| Clindamycin | 89 (16.3) | 120 (19.7) | 0.134 | 0.803 |
| Trimethoprim-sulfamethoxazole | 49 (9.0) | 57 (9.4) | 0.822 | 1.000 |
| Tetracycline | 81 (14.9) | 109 (17.9) | 0.161 | 0.807 |
| MDR | 84 (15.4) | 107 (17.6) | 0.319 | 1.000 |
