## Supplementary Table S9. Distribution of antimicrobial resistance and multidrug resistance among Streptococcus pneumoniae serotypes isolated from child for "Pneumococcal nasopharyngeal carriage in children 2–60 months of age in Novi Sad (Serbia), October 2022–September 2026"

| **Serotype** | **Isolates n** | **Penicillin G non-susceptible** | | **Ampicillin resistant** | | **Erythromycin resistant** | | **Clindamycin resistant** | | **Trimethoprim-sulfamethoxazole resistant** | | **Tetracycline resistant** | | **MDR** | |
| --- | --- | --- | --- | --- | --- | --- | --- | --- | --- | --- | --- | --- | --- | --- | --- |
|  |  | n | % | n | % | n | % | n | % | n | % | n | % | n | % |
| 11A | 111 | 0 | 0.0 | 2 | 1.8 | 5 | 4.5 | 3 | 2.7 | 6 | 5.4 | 2 | 1.8 | 2 | 1.8 |
| 10A | 94 | 0 | 0.0 | 0 | 0.0 | 1 | 1.1 | 0 | 0.0 | 0 | 0.0 | 0 | 0.0 | 0 | 0.0 |
| 15A | 93 | 1 | 1.1 | 2 | 2.2 | 36 | 38.7 | 34 | 36.6 | 2 | 2.2 | 30 | 32.3 | 27 | 29.0 |
| 15B | 82 | 0 | 0.0 | 0 | 0.0 | 8 | 9.8 | 4 | 4.9 | 6 | 7.3 | 3 | 3.7 | 3 | 3.7 |
| 23A | 82 | 0 | 0.0 | 0 | 0.0 | 3 | 3.7 | 3 | 3.7 | 1 | 1.2 | 1 | 1.2 | 1 | 1.2 |
| 23B | 81 | 0 | 0.0 | 0 | 0.0 | 2 | 2.5 | 1 | 1.2 | 18 | 22.2 | 2 | 2.5 | 1 | 1.2 |
| 19A | 64 | 2 | 3.1 | 11 | 17.2 | 29 | 45.3 | 28 | 43.8 | 19 | 29.7 | 23 | 35.9 | 24 | 37.5 |
| 6B | 63 | 1 | 1.6 | 7 | 11.1 | 51 | 81.0 | 41 | 65.1 | 4 | 6.3 | 39 | 61.9 | 40 | 63.5 |
| 15C | 52 | 1 | 1.9 | 2 | 3.8 | 7 | 13.5 | 5 | 9.6 | 5 | 9.6 | 4 | 7.7 | 5 | 9.6 |
| 21 | 51 | 1 | 2.0 | 3 | 5.9 | 9 | 17.6 | 5 | 9.8 | 3 | 5.9 | 8 | 15.7 | 7 | 13.7 |
| 24F | 48 | 3 | 6.3 | 0 | 0.0 | 36 | 75.0 | 37 | 77.1 | 12 | 25.0 | 29 | 60.4 | 31 | 64.6 |
| 35B | 42 | 1 | 2.4 | 5 | 11.9 | 9 | 21.4 | 0 | 0.0 | 2 | 4.8 | 0 | 0.0 | 1 | 2.4 |
| 6C | 42 | 0 | 0.0 | 1 | 2.4 | 35 | 83.3 | 30 | 71.4 | 3 | 7.1 | 28 | 66.7 | 28 | 66.7 |
| 3 | 32 | 0 | 0.0 | 0 | 0.0 | 1 | 3.1 | 1 | 3.1 | 0 | 0.0 | 1 | 3.1 | 1 | 3.1 |
| 35F | 29 | 0 | 0.0 | 0 | 0.0 | 2 | 6.9 | 1 | 3.4 | 1 | 3.4 | 1 | 3.4 | 1 | 3.4 |
| Others | 187 | 1 | 0.5 | 13 | 7.0 | 37 | 19.8 | 16 | 8.6 | 24 | 12.8 | 19 | 10.2 | 19 | 10.2 |
| Non typeable | 159 | 6 | 3.8 | 27 | 17.0 | 85 | 53.5 | 52 | 32.7 | 30 | 18.9 | 48 | 30.2 | 47 | 29.6 |
