## Supplemental Data 1 for "Pneumococcal nasopharyngeal carriage in children 2–60 months of age in Novi Sad (Serbia), October 2022–September 2026"

**Supplementary Table S10. Distribution of pneumococcal vaccine-type isolates according to vaccination status among children aged 2–60 months, Novi Sad, Serbia, 2022–2026.**

| Vaccination status | Isolates n | PCV10-type n (%) | PCV13-type n (%) | PCV15-type n (%) | PCV20-type n (%) | PCV20-NVT n (%) |
| --- | --- | --- | --- | --- | --- | --- |
| Fully vaccinated | 745 | 57 (7.7) | 125 (16.8) | 147 (19.7) | 358 (48.1) | 387 (51.9) |
| Partially vaccinated | 366 | 32 (8.7) | 72 (19.7) | 79 (21.6) | 172 (47.0) | 194 (53.0) |
| Unvaccinated | 42 | 5 (11.9) | 5 (11.9) | 5(11.9) | 15 (35.7) | 27 (64.3) |

Data are presented as numbers and percentages of pneumococcal isolates within each vaccination category. Repeated isolates from the same child were included; therefore, observations could not be assumed to be independent. The analysis was descriptive, and no statistical significance testing was performed, Vaccine-type categories represent cumulative serotype coverage of the respective PCV formulations, PCV20-NVT denotes isolates not belonging to PCV20 vaccine serotypes.
